# A study of gene expression in the living human brain

**DOI:** 10.1101/2023.04.21.23288916

**Authors:** Lora E. Liharska, You Jeong Park, Kimia Ziafat, Lillian Wilkins, Hannah Silk, Lisa M. Linares, Ryan C. Thompson, Eric Vornholt, Brendan Sullivan, Vanessa Cohen, Prashant Kota, Claudia Feng, Esther Cheng, Jessica S. Johnson, Marysia-Kolbe Rieder, Jia Huang, Joseph Scarpa, Jairo Polanco, Emily Moya, Alice Hashemi, Matthew A. Levin, Girish N. Nadkarni, Robert Sebra, John Crary, Eric E. Schadt, Noam D. Beckmann, Brian H. Kopell, Alexander W. Charney

**Affiliations:** Icahn School of Medicine at Mount Sinai

## Abstract

A goal of medical research is to determine the molecular basis of human brain health and illness. One way to achieve this goal is through observational studies of gene expression in human brain tissue. Due to the unavailability of brain tissue from living people, most such studies are performed using tissue from postmortem brain donors. An assumption underlying this practice is that gene expression in the postmortem human brain is an accurate representation of gene expression in the living human brain. Here, this assumption – which, until now, had not been adequately tested – is tested by comparing human prefrontal cortex gene expression between 275 living samples and 243 postmortem samples. Expression levels differed significantly for nearly 80% of genes, and a systematic examination of alternative explanations for this observation determined that these differences are not a consequence of cell type composition, RNA quality, postmortem interval, age, medication, morbidity, symptom severity, tissue pathology, sample handling, batch effects, or computational methods utilized. Analyses integrating the data generated for this study with data from earlier landmark studies that used tissue from postmortem brain donors showed that postmortem brain gene expression signatures of neurological and mental illnesses, as well as of normal traits such as aging, may not be accurate representations of these gene expression signatures in the living brain. By using tissue from large cohorts living people, future observational studies of human brain biology have the potential to (1) determine the medical research questions that can be addressed using postmortem tissue as a proxy for living tissue and (2) expand the scope of medical research to include questions about the molecular basis of human brain health and illness that can only be addressed in living people (e.g., “What happens at the molecular level in the brain as a person experiences an emotion?”).

## Introduction

A long-standing barrier to understanding the molecular basis of human brain health and illness is the unavailability of living human brain tissue for medical research^1^. With samples from living people largely unavailable, postmortem brain samples have become the standard tissue source for observational studies of human brain molecular biology, including studies of gene expression^2^. Accordingly, most gene expression studies of neurological and mental illnesses in humans have been performed using postmortem brain tissue, including landmark observational studies of schizophrenia (SCZ)^3^ and Alzheimer’s disease (ALZ)^4^. Since observational studies of gene expression in the postmortem human brain are often intended to discover the molecular biology underlying traits of the living human brain (e.g., neurodevelopment, aging, disease pathogenesis), they are often conducted based on the implicit assumption that gene expression in the postmortem brain is an accurate representation of gene expression in the living brain. Up until now, observational studies testing this assumption in humans have been small in scale, conducted prior to the advent of next-generation sequencing technologies, and limited to comparisons of living and postmortem cohorts not matched for key clinical and technical variables^5–8^. The Living Brain Project (LBP) developed a safe and scalable procedure to acquire prefrontal cortex (PFC) tissue from living people solely for medical research purposes (i.e., not for clinical purposes; ***Figure 1***). Here, in the flagship published report on samples obtained for the LBP, gene expression was compared between 275 PFC samples from living participants and 243 PFC samples from postmortem donors in order to test the assumption that gene expression in the postmortem brain is an accurate representation of gene expression in the living brain.

**Figure 1.**
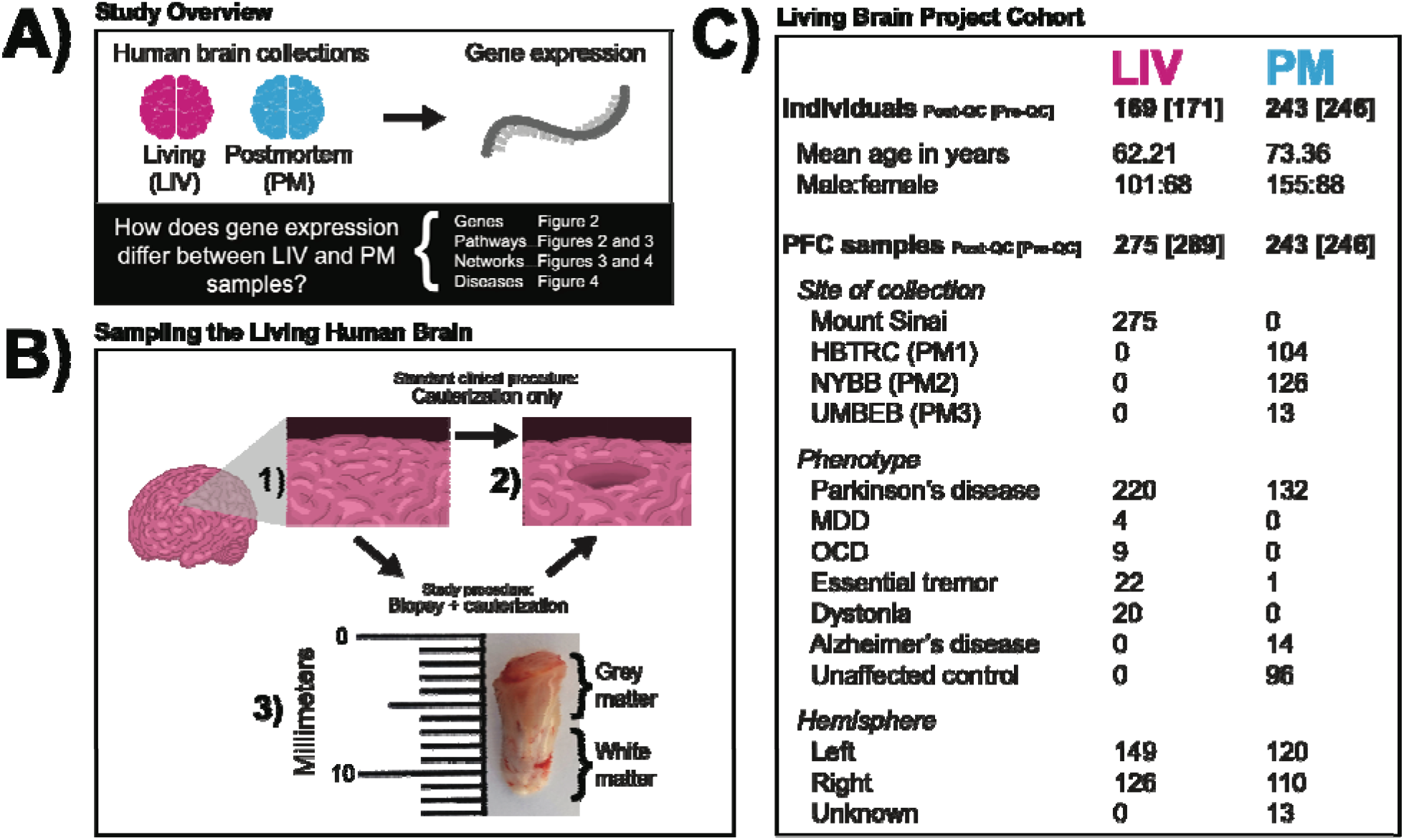
(A) Study Overview. The pink and blue colors represent LIV samples and PM samples, respectively, throughout all subsequent figures. (B) Sampling the Living Human Brain. The schematic illustrates that the standard clinical procedure and the modified study procedure amount to the same effective PFC volume loss. In 1) the state of the PFC at baseline is depicted. In 2) the final state of the PFC is depicted (i.e., the inner cylinder of tissue removed). The arrow and associated labels indicate how the final state of preparation is achieved in the standard clinical procedure (cauterization only) and the LBP procedure (biopsy followed by cauterization). Shown in 3) is a photograph of a PFC biopsy obtained for the LBP that is representative of the samples studied in this report. The top of the image is the anterior surface of the PFC. (C) Living Brain Project Cohort. Numbers refer to sample size (i.e., individuals or samples) except for age. The individual and sample numbers are identical for the PM cohort because only one sample was obtained per individual. Sample sizes inside and outside of the square brackets indicate counts prior to and after the application of quality control filters, respectively. [PFC = prefrontal cortex; QC = quality control; HBTRC (PM1) = Harvard Brain and Tissue Resource Center; NYBB (PM2) = New York Brain Bank at Columbia University; UMBEB (PM3) = University of Miami Brain Endowment Brain; MDD = major depressive disorder; OCD = obsessive-compulsive disorder]

## Results

### Living Brain Project cohort

PFC samples from living participants were obtained during neurosurgical procedures for deep brain stimulation (DBS), an elective treatment for neurological and mental illnesses such as Parkinson’s disease (PD), obsessive-compulsive disorder, essential tremor, and dystonia^9^. A common technique for safe implantation of the DBS electrode involves cauterizing a portion of the PFC, resulting in a small volume loss that has no discernible clinical consequence. For the LBP, a modification of this technique was developed to biopsy the PFC volume that would otherwise be cauterized (***Figure 1B***). Zero adverse events attributed to the biopsy procedure were observed over the study period. A total of 289 PFC biopsies (“LIV samples”) from 171 living participants were obtained during that time, including unilateral biopsies from 53 participants (40 from the left hemisphere and 13 from the right hemisphere) and bilateral biopsies from 118 participants. For comparison, a cohort of postmortem PFC samples (“PM samples”, N = 246) was assembled from three brain banks (“PM1”, N = 104; “PM2”, N = 129; “PM3”, N = 13). To the extent that it was possible, PM samples were matched to LIV samples for age and sex. The majority of samples were obtained from individuals with PD (***Figure 1C***), the most common indication for DBS.

All LIV samples and PM samples (N = 535) were processed for gene expression profiling together at the Icahn School of Medicine at Mount Sinai in New York City. RNA extraction, cDNA library preparation, and RNA sequencing were performed in batches constructed to contain approximately equal numbers of LIV samples and PM samples. After sequencing, raw nucleotide sequences (“reads”) were aligned to the reference transcriptome. Aligned reads were then counted and normalized for each gene in the reference. Genes with >=1 count per million reads in 10% of samples were retained for analysis (21,635 genes). Variables explaining unwanted variance in gene expression, including cell type composition and several technical sequencing metrics (***Supplementary Figures 1-4***), were identified through an adaptation of an iterative procedure established in previous RNA sequencing studies^10–12^ and included as covariates in subsequent statistical models. Excluded from analysis were samples with discordant labeled and genetically determined identities, samples unable to be aligned to the reference transcriptome for technical reasons, samples with low aligned read counts, and samples determined to be outliers for technical sequencing metrics. After completion of these quality control procedures, the cohort retained for analysis consisted of 518 samples (275 LIV samples and 243 PM samples) from 412 individuals (169 living participants and 243 postmortem donors) (***Figure 1C***). This cohort will be referred to as the “full LBP cohort.”

### Identifying the primary LIV-PM DE signature

In differential expression (DE) analysis, the expression level of every gene is tested for association with a trait of interest using a regression model. For a given gene, the regression model beta (by convention, the “logFC” value) for the trait of interest captures both the magnitude and direction of the gene-trait association. The set of gene-trait logFCs for all genes tested is collectively referred to as the “DE signature” of the trait, and genes with statistically significant associations with the trait are referred to as “differentially expressed genes” (DEGs). The trait of primary interest in the full LBP cohort was “LIV-PM status” (i.e., whether a PFC sample is from a living participant or a postmortem donor). DE of LIV-PM status (“LIV-PM DE”) was performed on 21,635 genes. The resulting LIV-PM DE signature (“the primary LIV-PM DE signature”) is provided in ***Supplementary Table 1***. Approximately 80% of genes were differentially expressed after adjusting for multiple tests using a false discovery rate (FDR) of 5% (17,186 “LIV-PM DEGs”, ***Figure 2A***), with 9,198 DEGs more highly expressed in LIV samples relative to PM samples (“LIV DEGs”; 42.51% of genes) and 7,988 DEGs more highly expressed in PM samples relative to LIV samples (“PM DEGs”; 36.92% of genes). The ability of gene expression to separate LIV samples from PM samples is visualized using principal components in ***Supplementary Figure 5***.

**Figure 2.**
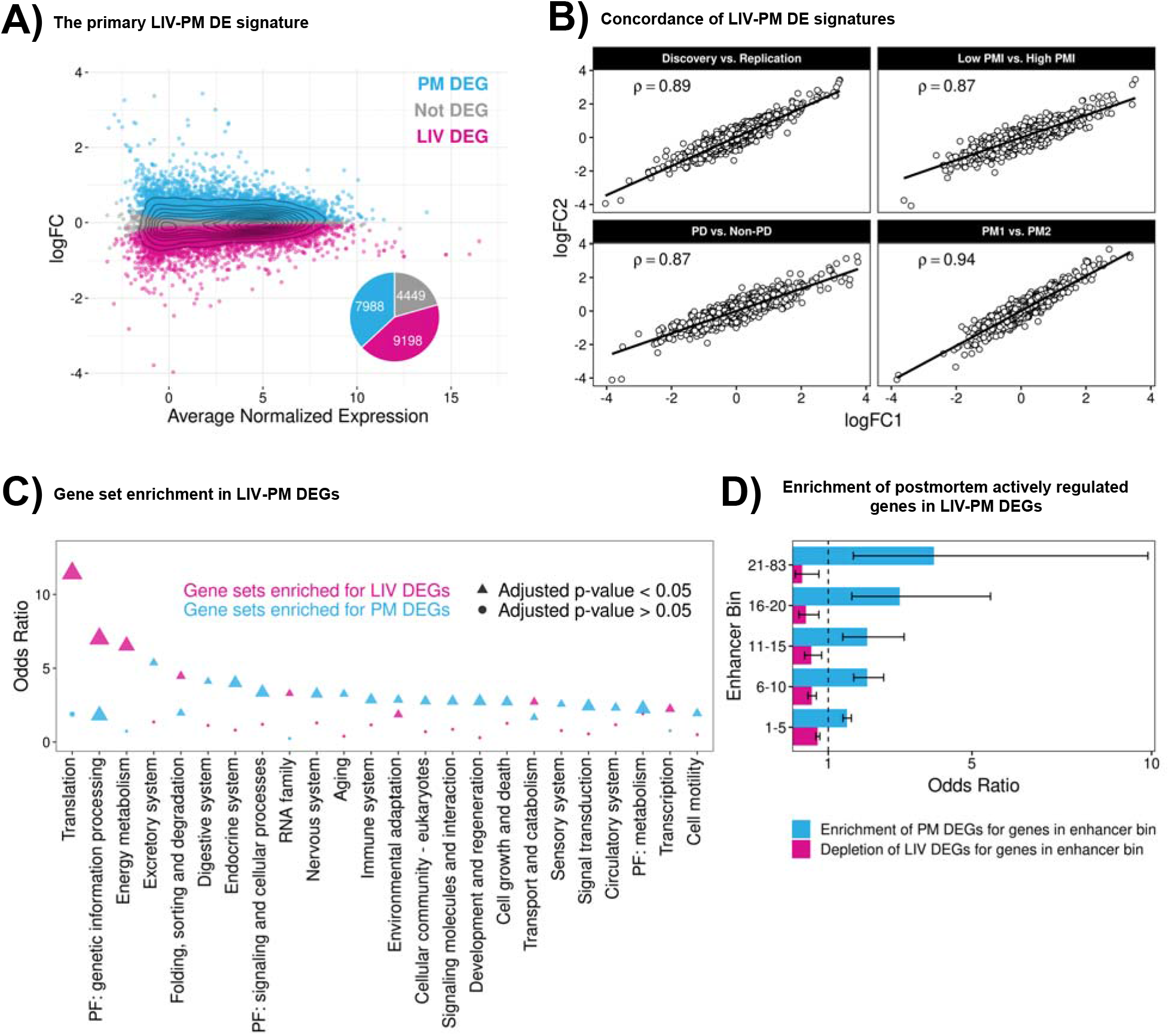
(A) LIV-PM DE results in the full LBP cohort. The scatterplot has one point for each of the 21,635 genes expressed in the full LBP cohort (275 LIV samples and 243 PM samples). The x-axis of the scatterplot is the average normalized expression level for each gene. The y-axis of the scatterplot is the LIV-PM DE regression model beta (by convention, the “logFC”) for each gene. Blue points represent DEGs upregulated in PM samples relative to LIV samples and pink points represent DEGs upregulated in LIV samples relative to PM samples. Grey points represent genes that were not differentially expressed between LIV samples and PM samples. The pie chart shows the fraction of scatterplot points that are blue, pink, and grey. The numbers in the pie chart indicate the number of LIV DEGs, PM DEGs, and Not DEGs. (B) LIV-PM DE Signature Comparisons. Each of the four scatterplots depict the relationship between LIV-PM DE signatures generated from independent subsets of LBP samples. X- and y-axes are the logFC values of the two LIV-PM DE signatures being compared in the plot. The Spearman’s correlation coefficient (ρ) between the two LIV-PM DE signatures is shown. The LIV-PM DE signatures compared in each plot are indicated by the plot title and fully described in the main text. For the “Low PMI vs High PMI” panel, only PM1 is depicted. (C) Gene set enrichment in LIV-PM DEGs. Each circle or triangle is a KEGG gene set tested for enrichment in LIV DEGs (pink circles and triangles) and PM DEGs (blue circles and triangles). The x-axis is the parent category of the gene set in the KEGG database. The y-axis is the Fisher’s Exact Test odds ratio for enrichment of a KEGG gene set in LIV DEGs and PM DEGs. Only parent categories with >=1 significant enrichment in either LIV DEGs or PM DEGs are included in the figure. For a given parent category, the KEGG gene set with the smallest p-value in LIV DEGs and PM DEGs is plotted. Triangles indicate that the enrichment test result is significant after p-value adjustment and circles indicate that the enrichment test result is not significant. The size of the triangles and circles corresponds to the -log_10_(p-value) of the Fisher’s Exact Test, meaning larger sizes are more statistically significant. The full KEGG gene set enrichment results for LIV DEGs and PM DEGs are provided in ***Supplementary Table 2*** with a column indicating the gene sets depicted in this figure. (D) Enrichment of postmortem actively regulated genes in LIV-PM DEGs. Each of the 3,852 actively regulated genes expressed in the full LBP cohort was assigned to one of five sequential “enhancer bins” based on the number of high-confidence active enhancers linked to the gene (i.e., 1-5, 6-10, 11-15, 16-20, or 21-83). The x-axis is the Fisher’s Exact Test odds ratio for the overlap of enhancer bin genes with LIV DEGs (pink bars) or PM DEGs (blue bars). The y-axis indicated the enhancer bin.

### Concordance of LIV-PM DE signatures

As postmortem samples are the standard tissue source used in gene expression studies of the human brain, the identification of the primary LIV-PM DE signature could have broad implications for neuroscience. Therefore, it is important to establish that the primary LIV-PM DE signature is not a consequence of confounding influences on (1) gene expression or (2) the ability to measure gene expression. Towards this end, analyses were performed for the following 14 potential confounding variables:

1. RNA sequencing wave
2. Institution of origin of the PM samples
3. Postmortem interval (PMI)
4. Diagnosis of PD in living participants and postmortem donors
5. Severity of PD symptoms in living participants
6. Dose of dopamine replacement therapy in living participants
7. Lewy body pathology in LIV and PM samples
8. Type and dose of anesthesia administered to living participants during DBS surgery
9. Method of LIV sample preservation upon collection during DBS surgery
10. RNA integrity number (RIN)
11. Age differences between living participants and postmortem donors
12. Threshold for defining lowly expressed genes
13. DE method
14. Cell type composition differences between LIV samples and PM samples.

This section is split into two parts – “Concordance of LIV-PM DE signatures: Part 1” and “Concordance of LIV-PM DE signatures: Part 2.” The 11 analyses performed in “Concordance of LIV-PM DE signatures: Part 1” used the same analytic framework, whereas the three analyses performed in “Concordance of LIV-PM DE signatures: Part 2” used slightly different analytic frameworks.

#### Concordance of LIV-PM DE signatures: Part 1

This part presents 11 out of the 14 analyses conducted to establish that the primary LIV-PM DE signature is a consequence of LIV-PM status and not of confounding influences on gene expression. For each of 11 potential confounding variables, a three-step analysis procedure was performed: (1) the full LBP cohort was stratified with respect to the potential confounding variable, (2) LIV-PM DE signatures were identified in the resulting groups, (3) the concordance of the LIV-PM DE signatures was assessed with respect to one another or to the primary LIV-PM DE signature.

##### 1. RNA sequencing wave

RNA sequencing of the full LBP cohort was conducted in two waves separated by over one year: the “discovery wave” and “replication wave.” The full LBP cohort was stratified into a discovery wave cohort (49 LIV samples and 57 PM samples) and a replication wave cohort (228 LIV samples and 187 PM samples not in the discovery wave cohort). LIV-PM DE was performed separately for the discovery wave cohort and the replication wave cohort. The concordance between the resulting LIV-PM DE signatures (Spearman’s ρ = 0.89, p-value < 2.2 x 10^-16^; ***Figure 2B***) suggests that the primary LIV-PM DE signature is not a consequence of RNA sequencing wave.

##### 2. Institution of origin of the PM samples

Institution of origin is a known potential confounder of postmortem brain gene expression^11^, and PM samples originated from three institutions (i.e., “brain banks”). LIV-PM DE was performed between PM1 samples and a random half of the LIV samples and between PM2 samples and the other half of the LIV samples. The concordance between the resulting LIV-PM DE signatures (Spearman’s ρ = 0.94, p-value < 2.2 x 10^-16^; ***Figure 2B***) suggests that the primary LIV-PM DE signature is not a consequence of brain bank.

##### 3. Postmortem interval (PMI)

Postmortem interval (PMI; i.e., the number of hours elapsed between death and postmortem tissue processing) is another known potential confounder of postmortem brain gene expression^13^. Since LIV samples, by definition, do not have a PMI, including PMI as a covariate in the LIV-PM DE regression model for each gene was not possible. PM1 samples and PM2 samples were stratified into “low PMI” and “high PMI” subsets, which were defined as the top and bottom quartiles of the PMI distributions, respectively, in each cohort (PM1: mean PMI in low PMI subset [26 samples] = 13.11, mean PMI in high PMI subset [26 samples] = 27.23; PM2: mean PMI in low PMI subset [28 samples] = 2.15, mean PMI in high PMI subset [27 samples] = 14.33). The LIV samples for this analysis were randomly split into four nearly equally sized subsets (three subsets of 69 samples and one subset of 68). DE was performed between (1) a LIV sample subset and the low PMI subset of PM1 (“LIV-PM:*LowPMI1* DE”), (2) a LIV sample subset and the low PMI subset of PM2 (“LIV-PM:*LowPMI2* DE”), (3) a LIV sample subset and the high PMI subset of PM1 (“LIV-PM:*HighPMI1* DE”), and (4) a LIV sample subset and the high PMI subset of PM2 (“LIV-PM:*HighPMI2* DE”). The concordance between (a) the LIV-PM:*LowPMI1* DE and LIV-PM:*HighPMI1* DE signatures (Spearman’s ρ = 0.87, p-value < 2.2 x 10^-16^; ***Figure 2B),*** and (b) the LIV-PM:*LowPMI2* DE and LIV-PM:*HighPMI2* DE signatures (Spearman’s ρ = 0.89, p-value < 2.2 x 10^-16^; ***Figure 2B)*** suggests that the primary LIV-PM DE signature is not simply a recapitulation of the known effect of PMI on postmortem brain gene expression. This observation is consistent with a previous observation made by the Genotype-Tissue Expression (GTEx) Project using RNA sequencing data from living and postmortem blood samples^14^.

##### 4. Diagnosis of PD in living participants and postmortem donors

The percentage of living participants diagnosed with PD (78.10%) was higher than the percentage of postmortem donors diagnosed with PD (54.32%). The full LBP cohort was stratified into a PD cohort (220 “LIV PD samples” and 132 “PM PD samples”) and a non-PD cohort (55 “LIV non-PD samples” and 111 “PM non-PD samples”). LIV-PM DE was performed separately for the PD cohort and the non-PD cohort. The concordance between the resulting LIV-PM DE signatures (Spearman’s ρ = 0.87, p-value < 2 x 10^-16^; *Figure 2B*) suggests that the primary LIV-PM DE signature is not a consequence of the different percentages of living participants and postmortem donors diagnosed with PD.

##### 5. Severity of PD symptoms in living participants

LIV PD samples were obtained from individuals with varying levels of PD symptom severity while PM PD samples were obtained from individuals whose PD symptom severity at the time of death was not provided by the brain banks. Living individuals with PD (N = 107) were categorized by PD symptom severity using scores from the Unified Parkinson’s Disease Rating Scale Motor subscale (UPDRS-III). Symptom severity was measured when individuals were off dopamine replacement therapy prior to the first DBS surgery (median time between symptom severity measurement and first DBS surgery = 60 days). UPDRS-III scores less than 40 were defined as low and UPDRS-III scores greater than or equal to 40 were defined as high. LIV-PM DE was performed between (1) LIV samples from individuals with low UPDRS-III scores (N = 92 samples from 60 individuals) and a random half of the PM PD samples (N = 66; “LIV:*lowUPDRS*-PM DE”), and (2) LIV samples from individuals with high UPDRS-III scores (N = 83 samples from 47 individuals) and the other half of the PM PD samples (N = 66; “LIV:*highUPDRS*-PM DE”). The concordance between the primary LIV-PM DE signature and (1) the LIV:*lowUPDRS*-PM DE signature (Spearman’s ρ = 0.95, p-value < 2.2 x 10^-16^), and (2) the LIV:*highUPDRS*-PM DE signature (Spearman’s ρ = 0.97, p-value < 2.2 x 10^-16^) suggests that the primary LIV-PM DE signature is not a consequence of PD symptom severity in the living cohort.

##### 6. Dose of dopamine replacement therapy in living participants

LIV PD samples were obtained from individuals chronically receiving dopamine replacement therapy (i.e., levodopa) while PM PD samples were obtained from individuals whose dopamine replacement therapy status at the time of death was not provided by the brain banks. LIV PD samples in this analysis were limited to 216 LIV PD samples (from 130 individuals). Levodopa dose (i.e., the dose of levodopa taken by a living participant at a time point close to LIV PD sample collection) less than 900 mg was defined as low and levodopa dose greater than or equal to 900 mg was defined as high. LIV-PM DE was performed between (1) LIV PD samples with low levodopa dose (N = 104) and a random half of the PM samples (N = 121; “LIV:*lowDOPA*-PM DE”), and (2) LIV PD samples with high levodopa dose (N = 112) and the other half of the PM samples (N = 122; “LIV:*highDOPA*-PM DE”). The concordance between the primary LIV-PM DE signature and (1) the LIV:*lowDOPA*-PM DE signature (Spearman’s ρ = 0.99, p-value < 2.2 x 10^-16^), and (2) the LIV:*highDOPA*-PM DE signature (Spearman’s ρ = 0.99, p-value < 2.2 x 10^-16^) suggests that the primary LIV-PM DE signature is not a consequence of levodopa dose.

##### 7. Lewy body pathology in LIV and PM samples

The living and postmortem cohorts included large proportions of individuals with PD at various stages of disease progression as measured on whole slide images by the density of intracellular aggregates of alpha-synuclein (i.e., Lewy bodies). LIV samples (N = 102) and PM samples (N = 93) were categorized as having:

1. No Lewy bodies (N = 24 LIV samples [i.e, the “LIV No” group]; N = 1 PM sample [i.e, the “PM No” group])
2. Low Lewy body density (N = 76 LIV samples [i.e, the “LIV Low” group]; N = 60 PM samples [i.e, the “PM Low” group])
3. Medium Lewy body density (N = 2 LIV samples [i.e, the “LIV Med” group]; N = 16 PM samples [i.e, the “PM Med” group])
4. High Lewy body density (N = 16 PM samples [i.e, the “PM High” group]).

After excluding the LIV Med and PM No groups due to small sample sizes, DE was performed between all pairs of LIV and PM groups, resulting in 10 DE signatures – six from DE of LIV-PM status (i.e., LIV*:No*-PM*:Low* DE, LIV*:No*-PM*:Med* DE, LIV*:No*-PM*:High* DE, LIV*:Low*-PM*:Low* DE, LIV*:Low*-PM*:Med* DE, LIV*:Low*-PM*:High* DE) and four DE signatures of Lewy body pathology (i.e., LIV*:No*-LIV*:Low* DE, PM*:Low*-PM*:Med* DE, PM*:Low*-PM*:High* DE, PM*:Med*-PM*:High* DE). The six DE signatures of LIV-PM status were highly correlated with the primary LIV-PM DE signature (Spearman’s ρ range = 0.89 – 0.97, all p-values < 2 x 10^-16^). In contrast, the four DE signatures of Lewy body pathology were much more lowly correlated with the primary LIV-PM DE signature (Spearman’s ρ range = −0.03 – 0.50, all p-values < 2 x 10^-16^). These findings suggest that the primary LIV-PM DE signature is (1) not a consequence of the density of Lewy bodies in LIV and PM samples, and (2) distinct from DE signatures of Lewy body pathology.

##### 8. Type and dose of anesthesia administered to living participants during DBS surgery

LIV samples were obtained from individuals under anesthesia during DBS surgery while PM samples were obtained from individuals whose anesthesia status at the time of death was not provided by the brain banks. Up to three types of anesthesia were given to each living individual: propofol, fentanyl, and dexmedetomidine. For the three types of anesthesia given, the following three-step procedure was performed:

1. LIV samples with available anesthesia data (N = 281) were split into quartiles representing four dose ranges of the anesthetic (N = 70 or N = 71 LIV samples in each quartile), then each quartile was subset for the samples in the full LBP cohort
2. PM samples in the full LBP cohort were randomly split into four groups (N = 60 or N = 61 PM samples in each group)
3. LIV-PM DE was performed between each quartile and one of the four PM groups.

Performing this procedure produced four LIV-PM DE signatures for each of the three types of anesthesia, for a total of 12 LIV-PM DE signatures (i.e., LIV*:PropofolQ1*-PM DE, LIV:*PropofolQ2*-PM DE, LIV:*PropofolQ3*-PM DE, LIV:*PropofolQ4*-PM DE, LIV*:DexmedetomidineQ1*-PM DE, LIV*:DexmedetomidineQ2*-PM DE, LIV*:DexmedetomidineQ3*-PM DE, LIV*:DexmedetomidineQ4*-PM DE, LIV*:FentanylQ1*-PM DE, LIV*:FentanylQ2*-PM DE, LIV:*FentanylQ3*-PM DE, LIV:*FentanylQ4*-PM DE). The concordance between these 12 LIV-PM DE signatures and the primary LIV-PM DE signature (Spearman’s ρ range = 0.97 – 0.98, all p-values < 2.2 x 10^-16^) suggests that the LIV-PM DE signature is not a consequence of the type and dose of anesthesia given to living individuals.

##### 9. Method of LIV sample preservation upon collection during DBS surgery

The method of preservation used for each LIV sample obtained during DBS surgery was either (1) placement in a tube of RNAlater or (2) placement in a tube on dry ice. DE was performed between (1) the LIV samples preserved by placement in RNAlater (N = 189) and a random half of the PM samples (N = 122; “LIV:*RNAlater*-PM DE”), and (2) the LIV samples preserved by placement on dry ice (N = 86) and the other half of the PM samples (N = 121; “LIV:*DryIce*-PM DE”). The concordance between the primary LIV-PM DE signature and (1) the LIV:*RNAlater*-PM DE signature (Spearman’s ρ = 0.98, p-value < 2.2 x 10^-16^), and (2) the LIV:*DryIce*-PM DE signature (Spearman’s ρ = 0.91, p-value < 2.2 x 10^-16^) suggests that the primary LIV-PM DE signature is not a consequence of the method of LIV sample preservation.

##### 10. RNA integrity number (RIN)

RIN, a known confounder of RNA sequencing data, was included as a covariate in the model used to discover the primary LIV-PM DE signature. This was done so that associations identified between LIV-PM status and gene expression levels are independent of associations between RIN and gene expression levels. RIN of LIV samples ranged from 5.5 to 8.8 and RIN of PM samples ranged from 4.1 to 9.7. LIV-PM DE additionally was performed between (1) the LIV samples with RIN less than or equal to 7 (N = 117) and PM samples with RIN less than or equal to 7 (N = 69; “LIV:*lowRIN*-PM:*lowRIN* DE”), and (2) the LIV samples with RIN greater than 7 (N = 158) and PM samples with RIN greater than 7 (N = 174; “LIV:*highRIN*-PM:*highRIN* DE”). The concordance between the primary LIV-PM DE signature and (1) the LIV:*lowRIN*-PM:*lowRIN* DE signature (Spearman’s ρ = 0.97, p-value < 2.2 x 10^-16^), and (2) the LIV:*highRIN*-PM:*highRIN* DE (Spearman’s ρ = 0.99, p-value < 2.2 x 10^-16^) suggests that the LIV-PM DE signature is not a consequence of RIN.

##### 11. Age differences between living participants and postmortem donors

Age at the time of tissue sampling is often accounted for as a covariate in DE analyses of human brain phenotypes due to the known relationship between age and human brain gene expression^15^. The iterative procedure employed for identification of confounder variables did not identify age as a covariate to be included in the model used to discover the primary LIV-PM DE signature. To ensure the primary LIV-PM DE signature is not a consequence of differences in the ages of living participants and postmortem donors, LIV-PM DE was performed between:

1. LIV samples with age less than 65 years (N = 151) and PM samples with age less than 65 years (N = 40; “LIV:*lowAge*-PM:*lowAge* DE”)
2. LIV samples with age greater than or equal to 65 years (N = 124) and PM samples with age greater than or equal to 65 years (N = 203; “LIV:*highAge*-PM:*highAge* DE”)
3. LIV samples with age less than 65 years and PM samples with age greater than or equal to 65 years (“LIV:*lowAge*-PM:*highAge* DE”)
4. LIV samples with age greater than or equal to 65 years and the PM samples with age less than 65 years (“LIV:*highAge*-PM:*lowAge* DE”).

The concordance between (1) the LIV:*lowAge*-PM:*lowAge* DE signature and the LIV:*highAge*-PM:*highAge* DE signature (Spearman’s ρ = 0.91, p-value < 2.2 x 10^-16^), (2) the LIV:*lowAge*-PM:*lowAge* DE signature and the LIV:*highAge*-PM:*lowAge* DE signature (Spearman’s ρ = 0.96, p-value < 2.2 x 10^-16^), (3) the LIV:*lowAge*-PM:*lowAge* DE signature and the LIV:*lowAge*-PM:*highAge* DE signature (Spearman’s ρ = 0.92, p-value < 2.2 x 10^-16^), (4) the LIV:*highAge*-PM:*highAge* DE signature and the LIV:*highAge*-PM:*lowAge* DE signature (Spearman’s ρ = 0.90, p-value < 2.2 x 10^-16^), (5) the LIV:*highAge*-PM:*highAge* DE signature and the LIV:*lowAge*-PM:*highAge* DE signature (Spearman’s ρ = 0.98, p-value < 2.2 x 10^-16^), and (6) the LIV:*highAge*-PM:*lowAge* DE signature and the LIV:*lowAge*-PM:*highAge* DE signature (Spearman’s ρ = 0.86, p-value < 2.2 x 10^-16^) suggests that the LIV-PM DE signature is not a consequence of age differences between living individuals and postmortem donors.

#### Concordance of LIV-PM DE signatures: Part 2

This part presents the final three of the 14 analyses conducted to establish that the primary LIV-PM DE signature is a consequence of LIV-PM status and not of confounding influences on gene expression.

##### 12. Threshold for defining lowly expressed genes

Following a standard practice for analysis of RNA sequencing data, in the current report genes retained for analysis (i.e., expressed genes) are defined as genes with at least 1 count per million in at least 10% of the LBP samples (“the 10% definition”). The LIV-PM DE signature observed using the 10% definition (i.e., the primary LIV-PM DE signature) contained 17,186 LIV-PM DEGs (79.4% of expressed genes). LIV-PM DE was additionally performed on the full LBP cohort with expressed genes defined as (1) genes with at least 1 count per million in at least 50% of samples (“the 50% definition”), and (2) genes with at least 1 count per million in at least 75% of samples (“the 75% definition”). The LIV-PM DE signatures observed using the 50% definition and the 75% definition contained 14,962 LIV-PM DEGs (80.3% of expressed genes) and 13,868 LIV-PM DEGs (80.9% of expressed genes), respectively. The concordance between the LIV-PM DE signature identified using the 10% definition and (1) the LIV-PM DE signature identified using the 50% definition (Spearman’s ρ = 0.99, p-value < 2 x 10^-16^), and (2) the LIV-PM DE signature identified using the 75% definition (Spearman’s ρ = 0.99, p-value < 2 x 10^-16^) suggests that the primary LIV-PM DE signature is not a consequence of the criteria used to define expressed genes.

##### 13. DE method

The primary LIV-PM DE signature was discovered using the dream() function of the variancePartition R package on a counts matrix normalized using the voomWithDreamWeights() function of the variancePartition R package. LIV-PM DE was additionally performed using the DESeq() function of the DESeq2 R package on a counts matrix consisting of 21,660 genes expressed in 169 LIV samples and 243 PM samples normalized using the DESeqDataSetFromMatrix() function of the DESeq2 R package (version 1.36.0)^16^. Using DESeq2, 74.2% of genes were identified as DEGs. The concordance between the primary LIV-PM DE signature and the DESeq2 LIV-PM DE signature (Spearman’s ρ = 0.96, p-value < 2.2 x 10^-16^) suggests that the primary LIV-PM DE signature is not a consequence of the software used to run DE.

##### 14. Cell type composition differences between LIV samples and PM samples

Cell fractions for the samples in the full LBP cohort were estimated using a cell type reference^17^ comprised of five cell types: glutamatergic neurons (GLU), GABA-ergic neurons (GABA), oligodendrocytes (ODC), astrocytes (AST), and microglia (MG). Neuronal cell fraction estimates were calculated by summing the GLU and GABA fraction estimates. Quality control procedures described in the methods section identified neuronal cell fraction as a covariate to be included in the model used to discover the primary LIV-PM DE signature. While neuronal fraction was the only cell fraction selected as a covariate in the model, an additional 13 possible combinations of cell fractions could have theoretically been used in the model (i.e., all combinations noted in the table below). To further establish that the primary LIV-PM DE signature is not a consequence of cell type composition, LIV-PM DE was performed for 12 of these 13 possible combinations of cell fractions (for one combination – neuronal, ODC, and AST – it was not possible to perform DE due to the high correlations between the cell fraction variables). The concordance between the primary LIV-PM DE signature and the resulting 12 LIV-PM DE signatures, presented in the following table, suggests that the primary LIV-PM DE signature is not a consequence of cell type composition. The table column descriptions are:

● “Model” – the formula in the methods section used to identify the LIV-PM DE signature reported in the row
● “Cell fractions” – the cell fractions included as covariates in the model used to identify the LIV-PM DE signature reported in the row
● “Concordance” – the Spearman’s correlation coefficient (ρ) between the primary LIV-PM DE signature and the LIV-PM DE signature reported in the row
● “Fraction DEG” – the fraction of 21,635 genes expressed in the full LBP cohort that were identified as LIV-PM DEGs in the LIV-PM DE signature reported in the row.

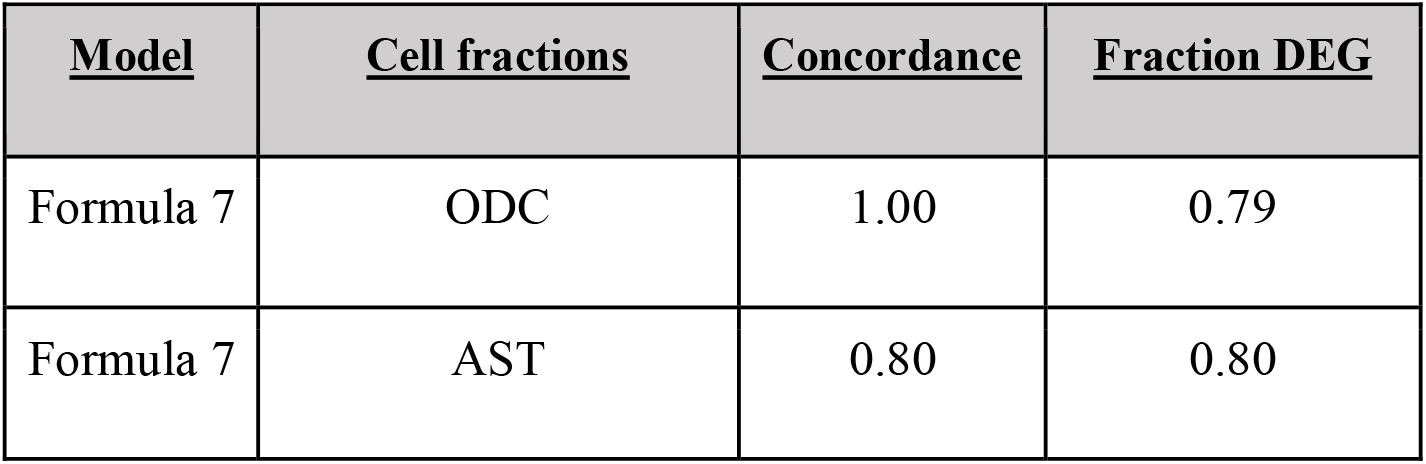
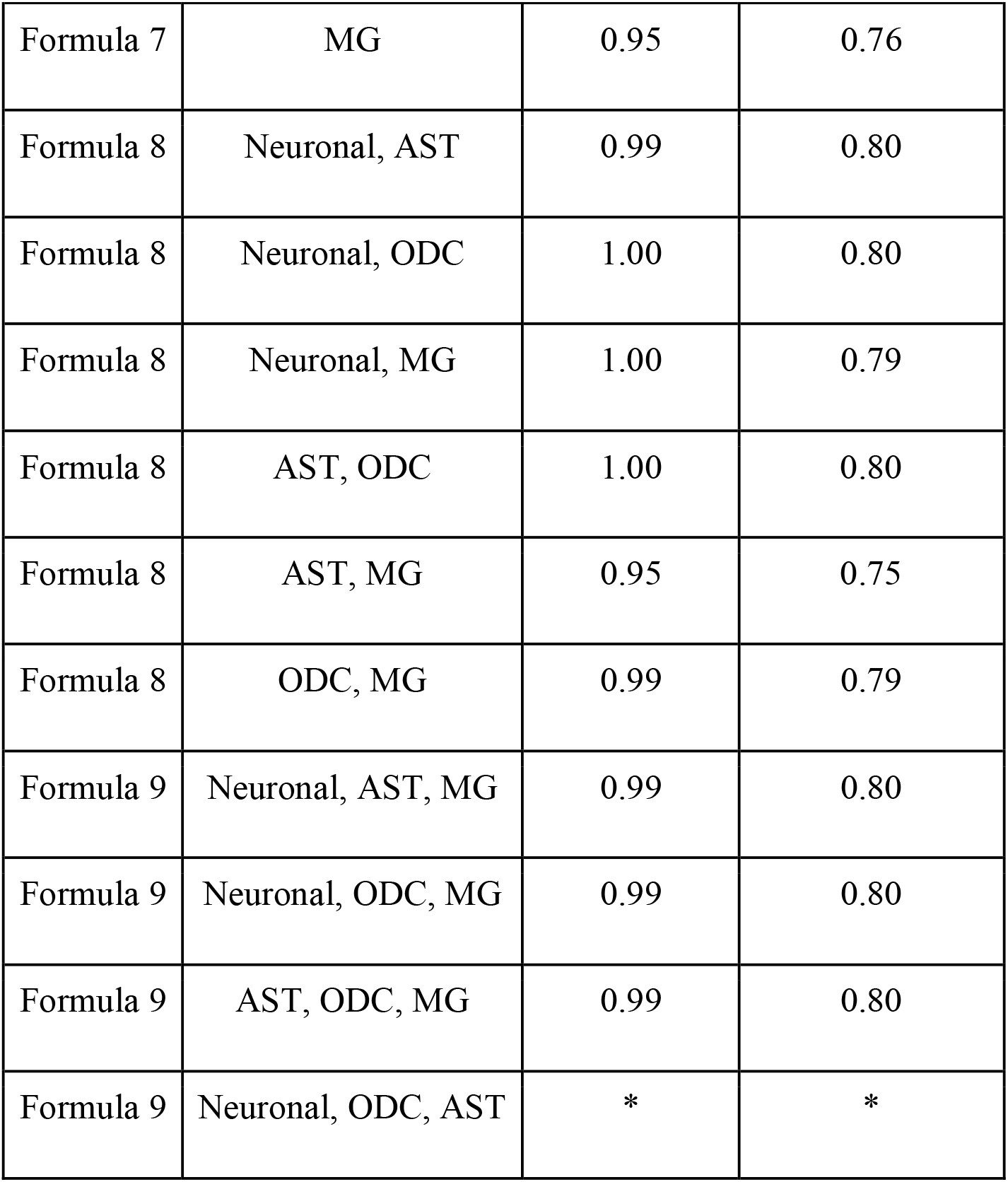

Together, the 14 analyses presented in this section suggest the primary LIV-PM DE signature is not a consequence of cell type composition, RNA quality, postmortem interval, age, medication, morbidity, symptom severity, tissue pathology, sample handling, batch effects, or computational methods utilized. The systematic exclusion of these alternative explanations for the primary LIV-PM DE signature leads to the conclusion that the primary LIV-PM DE signature is a consequence of LIV-PM status.

### Biological annotation of LIV-PM DE signature

Having concluded that the primary LIV-PM DE signature is a consequence of LIV-PM status, analyses were performed to elucidate the biological processes affected by LIV-PM status.

Housekeeping genes are genes that must be continuously expressed for cells to live^18^. To determine whether the expression of housekeeping genes differs between LIV samples and PM samples, LIV DEGs (N = 9,198) and PM DEGs (N = 7,988) were each tested for enrichment of housekeeping genes (N = 3,539)^19^ using Fisher’s exact test. LIV DEGs significantly overlapped with housekeeping genes (OR = 1.25, p-value = 1.66 x 10^-9^), while PM DEGs did not (OR = 1.06, p-value > 0.05). The higher expression of housekeeping genes in LIV samples compared to PM samples suggests that biological processes may be broadly affected by LIV-PM status. To verify this, LIV-PM DEGs were tested for enrichment of biological processes defined in the Kyoto Encyclopedia of Genes and Genomes (KEGG) database – which groups genes into sets based on curated annotations from public resources and published literature^20^. LIV DEGs and PM DEGs were enriched for 10 and 77 KEGG gene sets, respectively, after accounting for the 278 KEGG gene sets tested using an FDR of 5%. The full KEGG gene set results for LIV DEGs and PM DEGs are provided in ***Supplementary Table 2*** and a subset of these results are plotted in ***Figure 2C***. KEGG gene sets enriched in LIV DEGs were primarily related to protein translation and cellular respiration. PM DEGs, in contrast, were enriched for many diverse KEGG gene sets (e.g., ‘axon guidance,’ ‘chemokine signaling pathway,’ ‘carbohydrate digestion and absorption’) and the ‘transcription factor’ KEGG gene set was the most significantly enriched (Fisher’s exact test OR = 1.80, adjusted p-value = 1.18 x 10^-16^). None of the KEGG gene sets enriched in PM DEGs were enriched in LIV DEGs.

The enrichment of PM DEGs for transcription factors and many diverse biological processes suggests that gene expression regulation in the postmortem brain is contributing to the LIV-PM DE signature. Enhancers are short DNA sequences (50-1500 base pairs in length) that regulate gene expression by serving as binding sites for transcription factors to facilitate the transcription of associated genes. The PsychENCODE Consortium (PEC) has created a reference of “active enhancers” (i.e., enhancers located in regions of open chromatin) and “actively regulated genes” (i.e., genes linked to at least one active enhancer) in postmortem human brain samples^21^. Furthermore, the PEC has found that increased expression of an actively regulated gene is associated with higher numbers of active enhancers linked to the gene^21^. “High-confidence active enhancers” (i.e., active enhancers confirmed by multiple lines of evidence) were linked by PEC to 6,324 actively regulated genes^21^, of which 3,852 were expressed in the full LBP cohort. Fisher’s exact tests showed that these 3,852 actively regulated genes were significantly enriched in PM DEGs (OR = 1.61; adjusted one-sided p-value = 3.32 x 10^-39^) and significantly depleted of LIV DEGs (OR = 0.67; adjusted one-sided p-value = 4.40 x 10^-27^). Each of the 3,852 actively regulated genes expressed in the full LBP cohort was assigned to one of five sequential “enhancer bins” based on the number of high-confidence active enhancers linked to the gene (i.e., 1-5, 6-10, 11-15, 16-20, or 21-83). Fisher’s exact tests were performed between the actively regulated genes in each sequential enhancer bin and LIV-PM DEGs. The enrichment of PM DEGs and depletion of LIV DEGs in actively regulated genes became more pronounced as the number of high-confidence active enhancers linked to actively regulated genes increased (i.e., with each sequential enhancer bin) (***Figure 2D***). Using independent external data, these findings suggest that gene expression regulation in the postmortem brain is contributing to the LIV-PM DE signature.

### Gene-gene relationships in LIV samples and PM samples

Human brain health depends not only on the expression levels of individual genes but also on the relative expression levels of genes with respect to one another (“gene-gene relationships”). Based on the observation that 80% of genes expressed in the full LBP cohort are LIV-PM DEGs, the hypothesis that gene-gene relationships in the living human brain differ from gene-gene relationships in the postmortem human brain was tested. Pearson’s correlation coefficients were calculated for 234,025,795 gene-gene pairs (i.e., every combination of size 2 possible from 21,635 genes) in LIV samples and in PM samples. The LIV sample gene-gene correlations were subtracted from the PM sample gene-gene correlations and transformed to absolute values, resulting in the “LIV-PM correlation difference matrix.” The mean of this matrix (μ = 0.14) summarizes the extent to which gene-gene relationships differ between LIV samples and PM samples. To determine the statistical significance of this value, a null distribution of the means of 10,000 correlation difference matrices was generated by a random sampling procedure carried out on the full LBP cohort. The mean of the LIV-PM correlation difference matrix was greater than every value in the null distribution (empirical p-value < 0.0001; ***Figure 3A***), suggesting that gene-gene relationships throughout the transcriptome differ between LIV samples and PM samples in the full LBP cohort.

**Figure 3.**
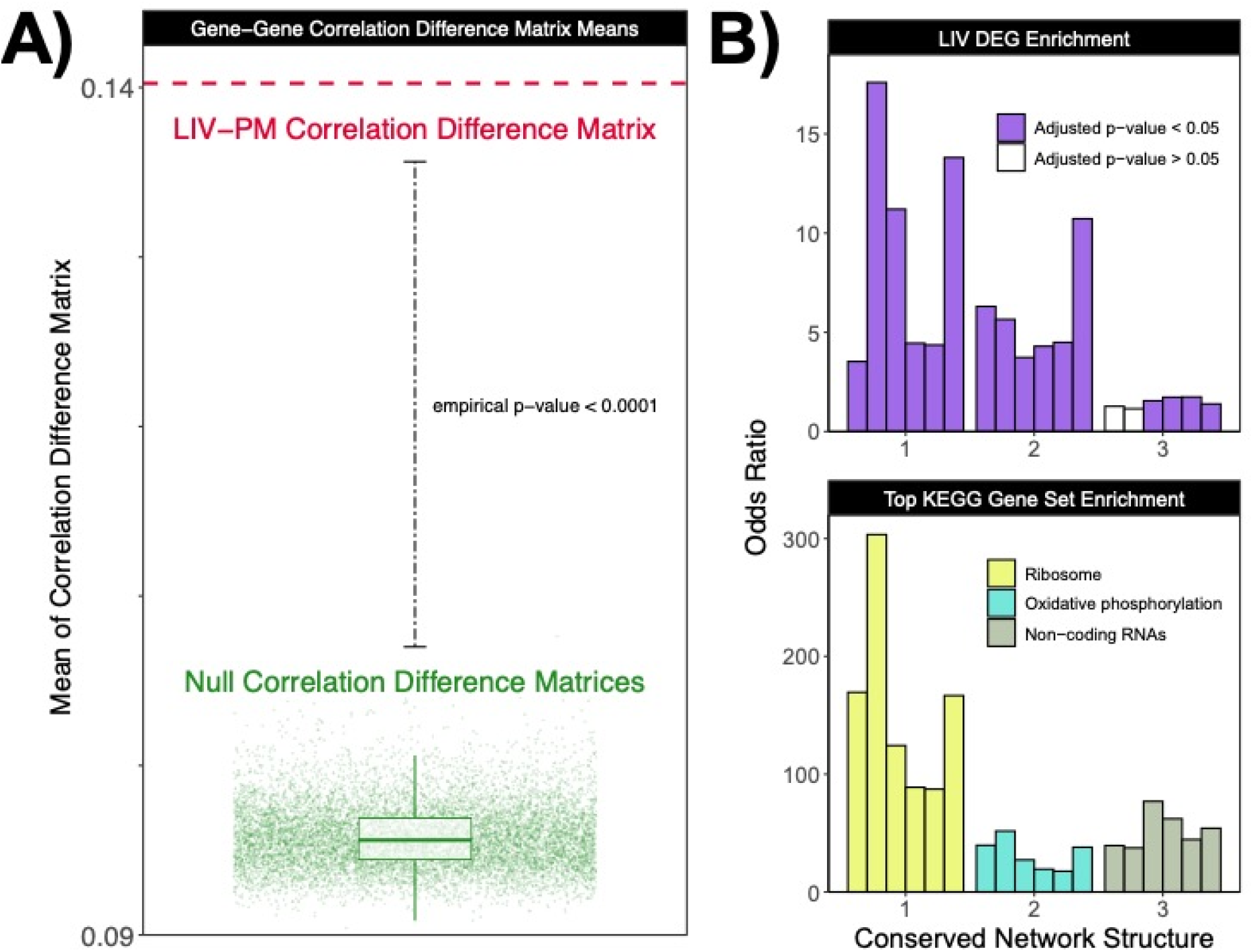
(A) Gene-gene Correlation Difference Matrix Means. The y-axis is the mean of each correlation difference matrix. Each of the 10,000 green points represents the mean of a “null correlation difference matrix” generated by comparing two random sample subsets. The overlaying boxplot summarizes the distribution of green points. The dotted red line represents the mean of the “LIV-PM correlation difference matrix” generated by comparing LIV samples to PM samples. (B) Conserved structures in LIV and PM networks. In both panels, the x-axis depicts the three conserved network structures identified across the six LBP networks that were enriched for LIV DEGs. The six modules of each conserved network structure are represented by bars. Each module is a network-specific representation of the conserved network structure, and the order of the bars for each conserved network structure is PM2 non-PD, PM2 PD, PM1 non-PD, PM1 PD, LIV non-PD, and LIV PD. The y-axis in the top panel is the odds ratio from the Fisher’s Exact Test of the enrichment of genes in the module for LIV DEGs. Purple bars indicate the enrichment test was significant after multiple testing correction. The y-axis in the bottom panel is the odds ratio from the Fisher’s Exact Test of the enrichment of genes in the module for the KEGG gene set most strongly associated with the module, and the color of the bars indicates the name of the KEGG gene set (all of the enrichment tests in the bottom panel were significant after multiple testing correction).

Despite these gene-gene relationship differences throughout the transcriptome, some of the 234,025,795 values in the LIV-PM correlation difference matrix were close to 0 (data not shown), implying that some organizational features of the human brain transcriptome may be conserved between living and postmortem tissues. To identify these features, a procedure was developed to compare living and postmortem human brain “co-expression networks,” which organize the transcriptome into groups of correlated genes (“modules”) that represent biological processes. Co-expression networks were constructed separately for LIV samples and for PM samples using weighted gene co-expression network analysis (WGCNA)^22^. Two LIV networks (i.e., networks constructed from LIV samples) and four PM networks (i.e., networks constructed from PM samples) were constructed from the full LBP cohort: LIV PD network (73 modules), LIV non-PD network (66 modules), PM1 PD network (92 modules), PM2 PD network (73 modules), PM1 non-PD network (79 modules), and PM2 non-PD network (63 modules). The six networks are provided in ***Supplementary Table 3***. All of the modules in every network were compared to all of the modules in the other networks to generate “module-module similarity metrics.” Employing a rule-based algorithm that takes as input the module-module similarity metrics for a set of networks, five “conserved network structures” were identified, each composed of six individual modules (i.e., one from each of the six networks) considered by the algorithm to be network-specific representations of the same biological processes (***Supplementary Table 4***). Conceptually, these five conserved network structures are organizational features of the brain transcriptome shared by LIV samples and PM samples in the full LBP cohort.

Given that 80% of all genes expressed in the full LBP cohort are LIV-PM DEGs, it was hypothesized that gene expression levels within the five conserved network structures would differ between LIV samples and PM samples. To test this hypothesis, the percentage of genes that are LIV-PM DEGs was calculated in each of the six network-specific representations of the five conserved network structures. In all 30 network-specific representations, the majority of genes were LIV-PM DEGs. This observation confirmed that although some organizational features of the brain transcriptome are conserved between LIV samples and PM samples, the expression levels of most of the genes within these features significantly differ between LIV samples and PM samples. In 19 of the 30 network-specific representations, the percentage of genes that were LIV DEGs was greater than the transcriptome-wide percentage of LIV DEGs (i.e., 42.51%). This observation led to the hypothesis that conserved network structures represent biological processes that are active in living tissues. Conserved network structures were therefore tested for enrichment of (1) LIV-PM DEGs and (2) KEGG gene sets (***Supplementary Table 4***). Significant enrichment of LIV DEGs was observed for three of the five conserved network structures. None of the conserved network structures were enriched for PM DEGs. The three conserved network structures enriched for LIV DEGs were annotated to either protein translation, cellular respiration, or non-coding RNA KEGG gene sets (***Figure 3B***), supporting the hypothesis that conserved network structures represent biological processes that are active in living brain tissues.

The gene-gene relationships observed in the full LBP cohort and presented in this section suggest (1) gene-gene relationships significantly differ between LIV samples and PM samples, (2) some organizational features of the transcriptome are conserved between LIV samples and PM samples, (3) the conserved organizational features capture biological processes essential to life, and (4) the expression levels of the genes involved in these essential biological processes differ between LIV samples and PM samples.

### DE signatures of brain phenotypes are affected by LIV-PM status

Collectively, the observations presented thus far suggest that gene expression in the postmortem human brain may not be an accurate representation of gene expression in the living human brain. This raises the possibility that disease DE signatures from postmortem studies of neurological and mental illnesses in humans may not be accurate representations of these gene expression signatures in the living brain. To investigate this possibility, the LIV-PM DE signature was compared to postmortem ALZ, SCZ, and PD DE signatures (***Figure 4A***). From publicly accessible data repositories, two cohorts each were obtained for ALZ^11^ and SCZ^3^: a discovery cohort (“ALZ1” [N = 246] and “SCZ1” [N = 306]) and a replication cohort (“ALZ2” [N = 343] and “SCZ2” [N = 414]). For PD, the discovery cohort (“PD1”) was defined as the PD cases and controls in PM1, and the replication cohort (“PD2”) was defined as the PD cases and controls in PM2. Details on the case and control ascertainment strategies utilized for the ALZ, SCZ, and PD cohorts can be found in the methods section along with relevant literature citations. After applying the same quality control pipeline described above for the full LBP cohort to the ALZ and SCZ cohorts, DE analysis comparing cases to controls was performed in all six cohorts (the complete results can be found in ***Supplementary Table 5***). For each of the three diseases, the two DE signatures generated were concordant with one another (Spearman’s ρ: ALZ1/ALZ2 = 0.49, SCZ1/SCZ2 = 0.26, PD1/PD2 = 0.63; all p-values < 2.2 x 10^-16^), highlighting the reproducibility of the postmortem disease DE signatures. Significant correlations were observed between (1) the primary LIV-PM DE signature and the ALZ1 DE signature (Spearman’s ρ = 0.40, p-value < 2.2 x 10^-16^), (2) the primary LIV-PM DE signature and the SCZ1 DE signature (Spearman’s ρ = 0.20, p-value = 1.65 x 10^-155^), and (3) the LIV-PM DE signature derived from the PM2 samples and half of the LIV samples and the PD1 DE signature (Spearman’s ρ = 0.64, p-value < 2.2 x 10^-16^). These observations were replicated between (1) the primary LIV-PM DE signature and the ALZ2 DE signature (Spearman’s ρ = 0.27, p-value = 2.09 x 10^-293^), (2) the primary LIV-PM DE signature and the SCZ2 DE signature (Spearman’s ρ = 0.17, p-value = 1.58 x 10^-110^), and (3) the LIV-PM DE signature derived from the PM1 samples and half of the LIV samples and the PD2 DE signature (Spearman’s ρ = 0.39, p-value < 2.2 x 10^-16^). In all six disease cohorts, Fisher’s exact tests showed that the DEGs with higher expression in cases relative to controls (“case DEGs”) significantly overlapped with PM DEGs (i.e., genes with higher expression in PM samples relative to LIV samples) (ORs 1.97 - 9.56, adjusted p-values < 1.02 x 10^-26^) but not with LIV DEGs (i.e., genes with higher expression in LIV samples relative to PM samples). Conversely, ALZ, SCZ, and PD DEGs with higher expression in controls relative to cases (“control DEGs”) significantly overlapped with LIV DEGs (ORs 1.46 - 10.60, adjusted p-values < 4.28 x 10^-9^) but not with PM DEGs (***Figure 4A, Supplementary Table 6***).

**Figure 4.**
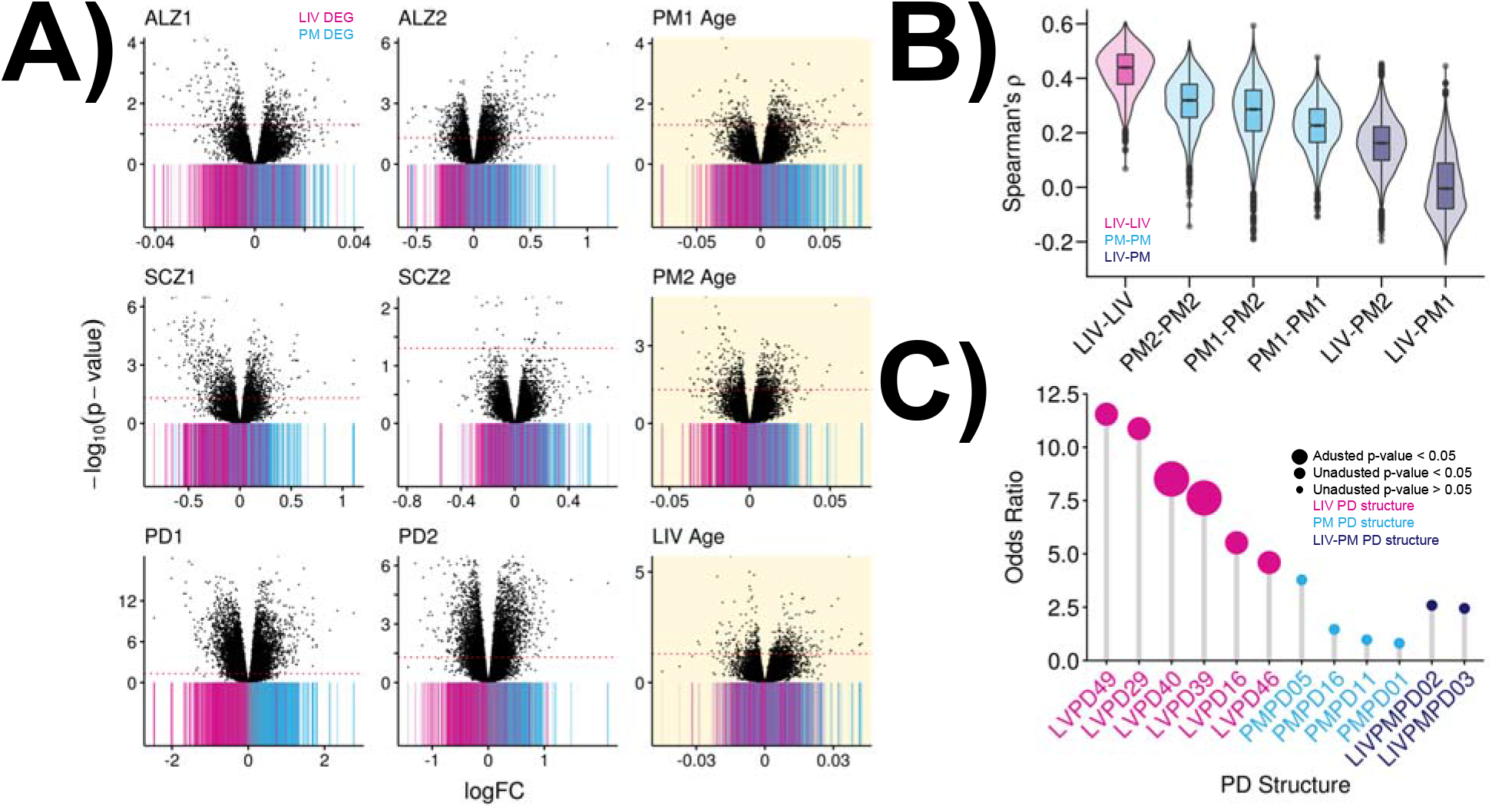
(A) Overlap between LIV-PM DE signature and ALZ, SCZ, PD, and Age DE signatures. Each scatterplot depicts the results of DE analysis for either ALZ (top row, columns 1 and 2), SCZ (middle row, columns 1 and 2), PD (bottom row, columns 1 and 2), or Age (column 3, highlighted in yellow). The x-axis of all nine scatterplots is the DE regression model beta (by convention, the “logFC”) for each gene. Positive logFC values indicate higher expression levels in cases (or with higher age) and negative logFC values indicate higher expression in controls (or with lower age). The y-axis is the -log_10_(p-value) for the DE test and the dotted red horizontal lines indicate the threshold for defining DEGs after multiple testing correction. ALZ, SCZ, PD, and PM Age DE signatures are derived from postmortem PFC samples. The LIV Age signature in the bottom row is from living PFC samples. The colored bar beneath every point in each scatterplot indicates whether the gene represented by the point is a LIV DEG (pink bar), PM DEG (blue bar), or not differentially expressed between LIV samples and PM samples (grey bars). (B) Stability of LIV and PM Age DE Signatures. The six violin plots depict distributions of correlations between Age DE signatures. The x-axis indicates the source of the samples used to generate the Age DE signatures compared. The colors indicate whether both sources are LIV samples (pink), both sources are PM samples (blue), or one source is LIV samples and the other is PM samples (purple). The Spearman’s correlation coefficient (ρ) between Age DE signatures is shown on the y-axis. (C) Enrichment of PD structures for PD GWAS genes. The y-axis is the Fisher’s Exact Test odds ratio for enrichment of PD GWAS genes in network-specific representations of each PD-specific conserved network structure on the x-axis (i.e., PD structure). The three classes of PD structures are shown: LIV PD structures (structures only seen in the LIV PD network; pink), PM PD structures (structures only seen in the two PM PD networks; blue), and LIV-PM PD structures (structures seen in LIV and PM PD networks; purple). For each structure, the network-specific representation with the highest odds ratio is shown. Only LIV PD structures that were enriched for PD GWAS genes (at the unadjusted or adjusted p-value threshold) are shown for that category. Since no PM PD structures and no LIV-PM PD structures were enriched for PD GWAS genes at the unadjusted p-value threshold, structures are shown for which all of the network-specific representations had an odds ratio > 0. The size of the circles corresponds to the significance of the enrichment test as indicated in the figure legend.

In order to ensure that the characteristic pattern of overlap observed between the LIV-PM DE signature and postmortem case-control DE signatures (i.e., case DEGs overlap PM DEGs and control DEGs overlap LIV DEGs) was not a consequence of the gene expression data processing procedures utilized in the current study, the overlap between the primary LIV-PM DE signature and published postmortem case-control DE signatures was assessed. The characteristic pattern of overlap was reproduced using published DE signatures of:

1. ALZ from a postmortem brain case-control study which included ALZ1 and ALZ2 cohorts plus additional ALZ case-control cohorts^23^ (case DEG enrichment for PM DEGs OR = 3.50, adjusted p-value = 6.53 x 10^-202^; control DEG enrichment for LIV DEGs OR = 3.99, adjusted p-value = 7.82 x 10^-261^)
2. SCZ from a postmortem brain case-control study which included SCZ1 and SCZ2 cohorts plus additional SCZ case-control cohorts^24^ (case DEG enrichment for PM DEGs = 3.72, 4.04 x 10^-175^; control DEG enrichment for LIV DEGs = 2.67, 3.43 x 10^-95^)
3. PD from a postmortem brain case-control study which did not include any samples from PD1 or PD2^25^ (case DEG enrichment for PM DEGs OR = 3.42, adjusted p-value = 3.08 x 10^-20^; control DEG enrichment for LIV DEGs = 1.54, adjusted p-value = 1.11 x 10^-7^)
4. bipolar disorder (BD) from a postmortem brain case-control study^24^ (case DEG enrichment for PM DEGs OR = 2.07, adjusted p-value = 1.41 x 10^-19^; control DEG enrichment for LIV DEGs = 2.34, adjusted p-value = 6.21 x 10^-14^)
5. autism spectrum disorder (ASD) from one postmortem brain case-control study^24^ (case DEG enrichment for PM DEGs OR = 3.80, adjusted p-value = 2.24 x 10^-61^; control DEG enrichment for LIV DEGs OR = 2.41, adjusted p-value = 7.79 x 10^-33^) and an additional 12 ASD DE signatures from a subsequent study^26^ (ORs and adjusted p-values in ***Supplementary Table 6***).

***Supplementary Table 6*** provides (1) the complete set of results establishing the characteristic pattern of overlap between the LIV-PM DE signature and postmortem case-control DE signatures and (2) overlap of pairs of postmortem case-control DEG sets (e.g., ALZ1 DEGs with SCZ1 DEGs).

The characteristic pattern of overlap between the LIV-PM DE signature and the postmortem ALZ, SCZ, PD, BD, and ASD DE signatures indirectly supports the hypothesis that for phenotypes related to human brain health, DE signatures identified in postmortem tissue may not always be accurate representations of molecular processes occurring in the living brain. To directly test this hypothesis, a phenotype is required for which DE signatures derived separately from LIV samples and PM samples can be compared. This could not be done for any of the case-control phenotypes studied in the current report due to the lack of required samples (i.e., PFC samples from living healthy controls, living ALZ cases, living SCZ cases, living BD cases, and living ASD cases). Age, however, is a phenotype with a known DE signature in the postmortem human brain^15^ for which DE could be performed separately in LIV samples and PM samples. To test whether the DE signature of human age changes based on the LIV-PM status of the brain samples analyzed, DE was performed in the full LBP cohort for the interaction between the age of the individual and the LIV-PM status of the PFC sample. At the current sample size, 707 DEGs were identified for this interaction variable. To estimate the proportion of genes that deviate from the null hypothesis that there is no relationship between gene expression and this interaction variable, the π_1_ statistic was used^27^. It was estimated that given sufficient samples, at least 35% of genes would be DEGs for the interaction variable. This suggests that the age DE signature derived from human brain samples changes depending on whether the samples are obtained from living individuals or postmortem donors.

To test whether DE signatures from samples obtained from living individuals and postmortem donors are equally accurate representations of the true DE signature for a phenotype (i.e., age), two approaches were taken. The first approach was to empirically evaluate the concordance of age DE signatures in LIV samples and in PM samples (“LIV Age DE signatures” and “PM Age DE signatures,” respectively), under the premise that concordance reflects true DE signal. In each of 2,000 permutations, six subsets of the full LBP cohort were created by randomly splitting in half the LIV samples, PM1 samples, and PM2 samples, respectively. Age DE signatures were generated for each of the six subsets, and pairwise correlations between the signatures were calculated. The correlations between pairs of LIV Age DE signatures were higher than the correlations between pairs of PM Age DE signatures, and this was observed regardless of the source of the PM Age DE signatures compared (i.e., PM1/PM1, PM2/PM2, PM1/PM2) (***Figure 4B***). Correlations were lowest between LIV Age DE signatures and PM Age DE signatures (***Figure 4B***), consistent with the results of the interaction variable DE analysis (i.e., the age DE signature varies with LIV-PM status). No significant association was observed between postmortem interval (PMI) and age in either PM1 samples (Pearson’s correlation coefficient = - 0.12, p-value = 0.22) or PM2 samples (Pearson’s correlation coefficient = −0.09, p-value = 0.35). This implies that even when accounting for potential confounders of postmortem gene expression, for a given trait, the DE signature in postmortem brain tissue may not always be an accurate representation of the DE signature in living brain tissue.

The second approach to test whether DE signatures from samples obtained from living individuals and postmortem donors are equally accurate representations of the true DE signature for a phenotype (i.e., age) was to evaluate whether DEGs from the LIV and PM age DE analyses (“LIV Age DEGs” and “PM Age DEGs”) overlap with DEGs for LIV-PM status in the manner characteristic of postmortem ALZ, SCZ, PD, BD, and ASD DEGs (i.e., case DEGs overlapping PM DEGs and control DEGs overlapping LIV DEGs; ***Figure 4A, Supplementary Table 6***). The premise behind this approach is that if the PM Age DEGs demonstrate the characteristic pattern of overlap and the LIV Age DEGs do not, it would suggest the postmortem state systematically confounds DE signatures of phenotypes related to human brain function. Age DEGs with a positive logFC (i.e., DEGs whose expression increases with age) are referred to as “Higher Age DEGs” and age DEGs with a negative logFC (i.e., DEGs whose expression decreases with age) are referred to as “Lower Age DEGs.” PM1 and PM2 Higher Age DEGs significantly overlapped with PM DEGs (ORs, adjusted p-values: PM1 = 9.40, 3.56 x 10^-68^; PM2 = 6.36, 7.71 x 10^-67^) but not with LIV DEGs (***Supplementary Table 6***). PM1 and PM2 Lower Age DEGs significantly overlapped with LIV DEGs (ORs, adjusted p-values: PM1 = 2.79, 2.84 x 10^-9^; PM2 = 5.05, 5.18 x 10^-47^) but not with PM DEGs (***Supplementary Table 6*)**. LIV Higher Age DEGs did not significantly overlap with PM DEGs or with LIV DEGs (***Supplementary Table 6*)**. LIV Lower Age DEGs significantly overlapped with LIV DEGs, though the effect size of the enrichment (OR = 1.43, adjusted p-value = 4.69 x 10^-3^) was less than half that seen in PM1 Lower Age DEGs and PM2 Lower Age DEGs (***Supplementary Table 6*)**, despite the LIV sample size being more than twice that of PM1 and PM2 (***Figure 1C***). LIV Lower Age DEGs did not overlap with PM DEGs (***Figure 4A*** and ***Supplementary Table 6)***. To assess the reproducibility of the overlap between LIV-PM DEGs and age DEGs from postmortem brain samples, DE for age was performed in the ALZ1, ALZ2, SCZ1, and SCZ2 cohorts, and the resulting age DE signatures were compared to the primary LIV-PM DE signature. For these analyses, the ALZ1, ALZ2, SCZ1, and SCZ2 cohorts were matched to the age ranges of the PM1 and PM2 samples (***Supplementary Figure 6***). The enrichments of Higher Age DEGs in PM DEGs and of Lower Age DEGs in LIV DEGs were replicated in the ALZ1, ALZ2, and SCZ2 cohorts (***Supplementary Table 6***).

In summary, the analyses presented in this section suggest that (1) disease DEGs identified from postmortem human brain tissues demonstrate a characteristic pattern of overlap with LIV-PM DEGs (i.e., case DEGs overlap PM DEGs and control DEGs overlap LIV DEGs), (2) the DE signature for age varies depending on whether LIV samples or PM samples are studied, (3) the concordance between DE signatures for age is highest when LIV samples are studied, and (4) the characteristic pattern of overlap observed between LIV-PM DEGs and age DEGs identified in PM samples (i.e., Higher Age DEGs overlap PM DEGs and Lower Age DEGs overlap LIV DEGs) is not observed for age DEGs identified in LIV samples. Together, these four findings suggest, but do not definitively prove, that postmortem brain gene expression signatures of neurological and mental illnesses, as well as of normal phenotypes such as aging, may not be accurate representations of these gene expression signatures in the living brain.

### PD pathogenesis is captured in living brain co-expression networks

Co-expression networks constructed from postmortem brain gene expression data are often leveraged to gain insights into the pathogenesis of neurological and mental illnesses^3,11,21,28–30^. As described above, it was not possible to derive PD DE signatures separately from LIV samples and PM samples due to the lack of PFC samples from living healthy controls. However, the presence of PFC samples from both living PD cases and postmortem PD cases allowed for LIV PD networks and PM PD networks to be constructed separately to test if one better captures the mechanisms underlying PD pathogenesis. Three sets of PD-specific conserved network structures were defined: “LIV PD structures” were defined as structures only seen in the LIV PD network; “PM PD structures” were defined as structures only seen in the two PM PD networks; and “LIV-PM PD structures” were defined as structures seen in all three PD networks. The set of genes used to represent PD pathogenesis was defined in a genome-wide association study (GWAS) of PD^31^ and Fisher’s exact tests were performed between these PD GWAS genes and the PD-specific conserved network structures. No PM PD structures or LIV-PM PD structures were enriched for PD GWAS genes at an unadjusted p-value threshold. In contrast, six LIV PD structures showed enrichment for PD GWAS genes using an unadjusted p-value threshold of 0.05 and two remained significant after multiple test correction (***Figure 4C***). Four of the six LIV PD structures enriched for PD GWAS genes were annotated to at least one KEGG gene set (***Supplementary Table 7***). One of the structures (LIVPD29) was annotated to the “synaptic vesicle cycle” KEGG gene set and included the gene *SNCA*, which codes for alpha-synuclein, a regulator of synaptic vesicle trafficking and the primary component of the neuropathological hallmark of PD – the Lewy body^32^. These findings provide evidence, but do not definitively prove, that the pathogenesis of PD may be better captured in gene expression data from living brain samples compared to postmortem brain samples.

## Discussion

With this report, the LBP makes three primary contributions to medical research. First, it describes a safe and scalable approach to sampling living human PFC tissue solely for medical research purposes (i.e., not for clinical purposes). Second, it presents multiple lines of evidence that gene expression in postmortem human brain tissue may not always be an accurate representation of gene expression in living human brain tissue. Third, it suggests that postmortem brain gene expression signatures of neurological and mental illnesses, as well as of normal phenotypes such as aging, may not always be accurate representations of these gene expression signatures in the living brain.

The findings of this report can be viewed with confidence based on (1) the strengths of the study design, (2) the systematic ruling out of alternative explanations for findings, and (3) the confirmation of findings using external data sources when possible. Strengths of the study design relative to earlier work on the topic^5–8^ include (a) a large increase in sample size, (b) the use of next-generation sequencing technology to quantify every gene expressed in the brain, (c) randomization to minimize batch effects at the key steps of the sequencing experiment (i.e., RNA extraction, cDNA library preparation, RNA sequencing), and (d) the public sharing of all data for the scientific community to interrogate. A systematic examination of alternative explanations for the differences in gene expression observed between LIV samples and PM samples ruled out the possibility that these differences are a consequence of cell type composition, RNA quality, postmortem interval, age, medication, morbidity, symptom severity, tissue pathology, sample handling, batch effects, or computational methods utilized. Analyses of external data sourced from a landmark postmortem human brain study showed that gene expression regulation in the postmortem brain is likely contributing to the differences in gene expression observed between living and postmortem brain tissues.

The study had notable limitations and leaves several key questions unresolved. The lack of PFC samples from living healthy controls, living ALZ cases, living SCZ cases, living BD cases, and living ASD cases prevented the direct comparison between gene expression signatures of disease identified in living human brain and in postmortem human brain. As the scope of the current study was limited to gene expression in bulk PFC samples, future studies should determine if differences between living and postmortem brain tissues are (1) present at other levels of molecular biology (e.g., protein expression) and (2) consistent across cell types and brain regions. Depending on the research question of interest, the molecular differences that exist between living and postmortem human brain tissues may or may not matter. Therefore, future work should aim to determine the research questions that can and cannot be answered using the postmortem human brain as a proxy for the living human brain at the molecular level.

The LBP, while limited to the context of DBS surgery at a single study site, provides a blueprint for integrating medical research activities (i.e., informed consent, sample collection, biobanking) into existing clinical workflows to acquire living human brain tissue. Several hundred PFC biopsies were safely obtained for the LBP solely for medical research purposes (i.e., not for clinical purposes) by making a simple modification to standard clinical practice (i.e., saving the PFC tissue that would otherwise be cauterized). Beyond DBS, similar integration of medical research activities into clinical workflows is likely possible for many neurosurgical procedures. Each year, over 10 million individuals undergo neurosurgery globally, many of whom have secondary neurologic or psychiatric diagnoses (i.e., diagnoses that are not the reason for the surgery)^33–36^. Obtaining brain biopsies for medical research from even a small fraction of these individuals would rapidly build biobanks of living brain tissue for studying the molecular foundations of a wide range of neurologic and psychiatric conditions.

At the time of writing, most gene expression studies of the human brain continue to only use postmortem tissue. The findings of this report suggest that brain tissue from living people should also be prioritized for medical research. Doing so will further elucidate the differences between living and postmortem tissue and allow for the development of statistical methods that account for these differences, thereby improving the ability of postmortem tissue to accurately represent living tissue. By providing the data needed for the development of these methods, studies such as the LBP should ultimately make postmortem human brain samples more – not less – valuable for advancing knowledge of the molecular basis of human brain health and illness. Prioritizing the study of living human brain tissue will also expand the scope of medical research to include questions regarding the molecular basis of human brain health and illness that (1) cannot be addressed in postmortem tissues due to the limited clinical data available on postmortem donors, and (2) can only be addressed in the context of studying living people (e.g., “What happens at the molecular level in the brain as a person experiences an emotion?”).

## Supplementary Figures

**Supplementary Figure 1:**
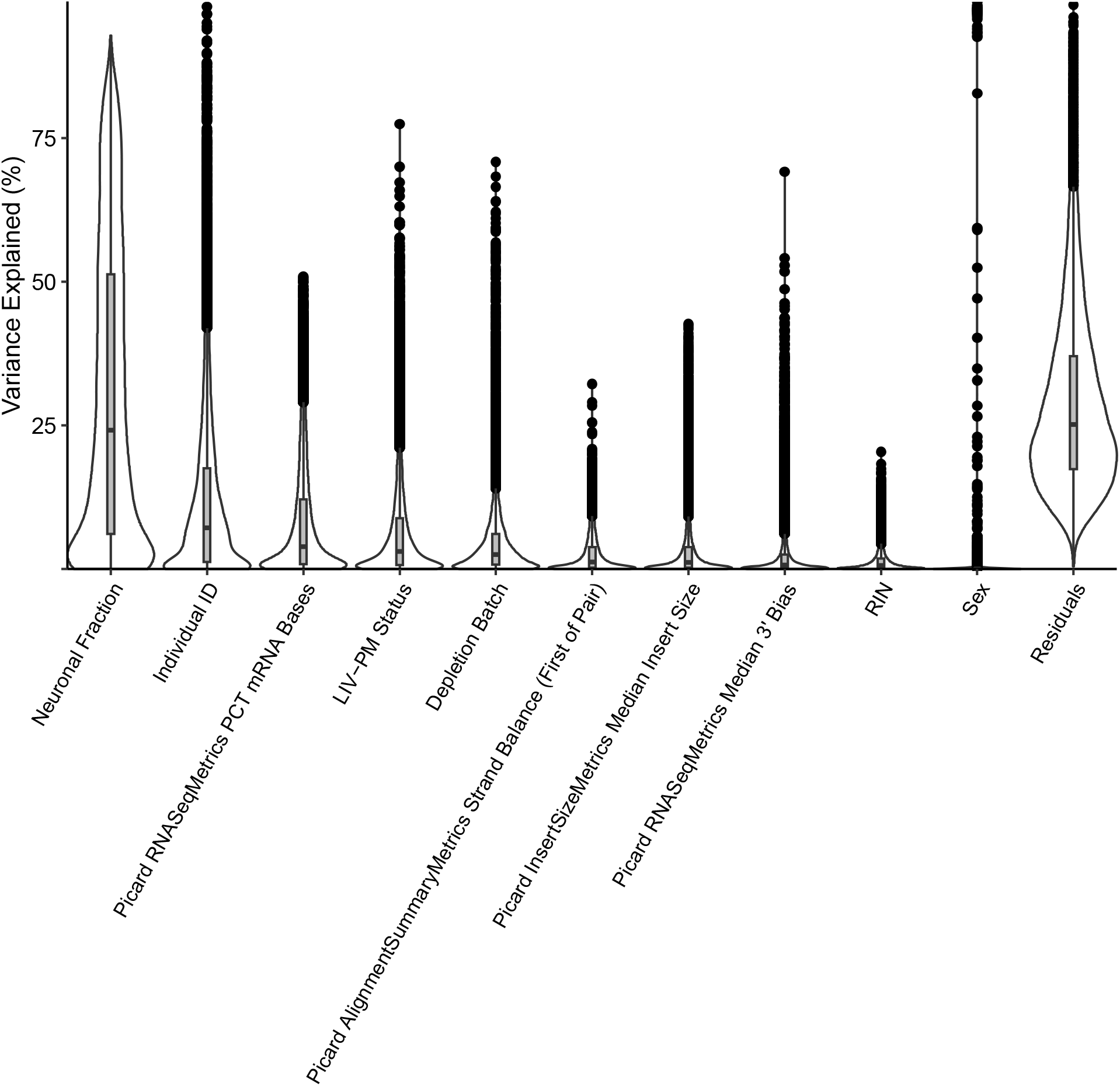
For each gene expressed in the full LBP cohort, the percent of variance in expression explained by each variable in the regression model (Formula XX) used to discover the primary LIV-PM DE signature was calculated. For each variable, the distribution of the resulting percentages is shown using the default parameters of the plotVarPart() function from the variancePartition R package.

**Supplementary Figure 2:**
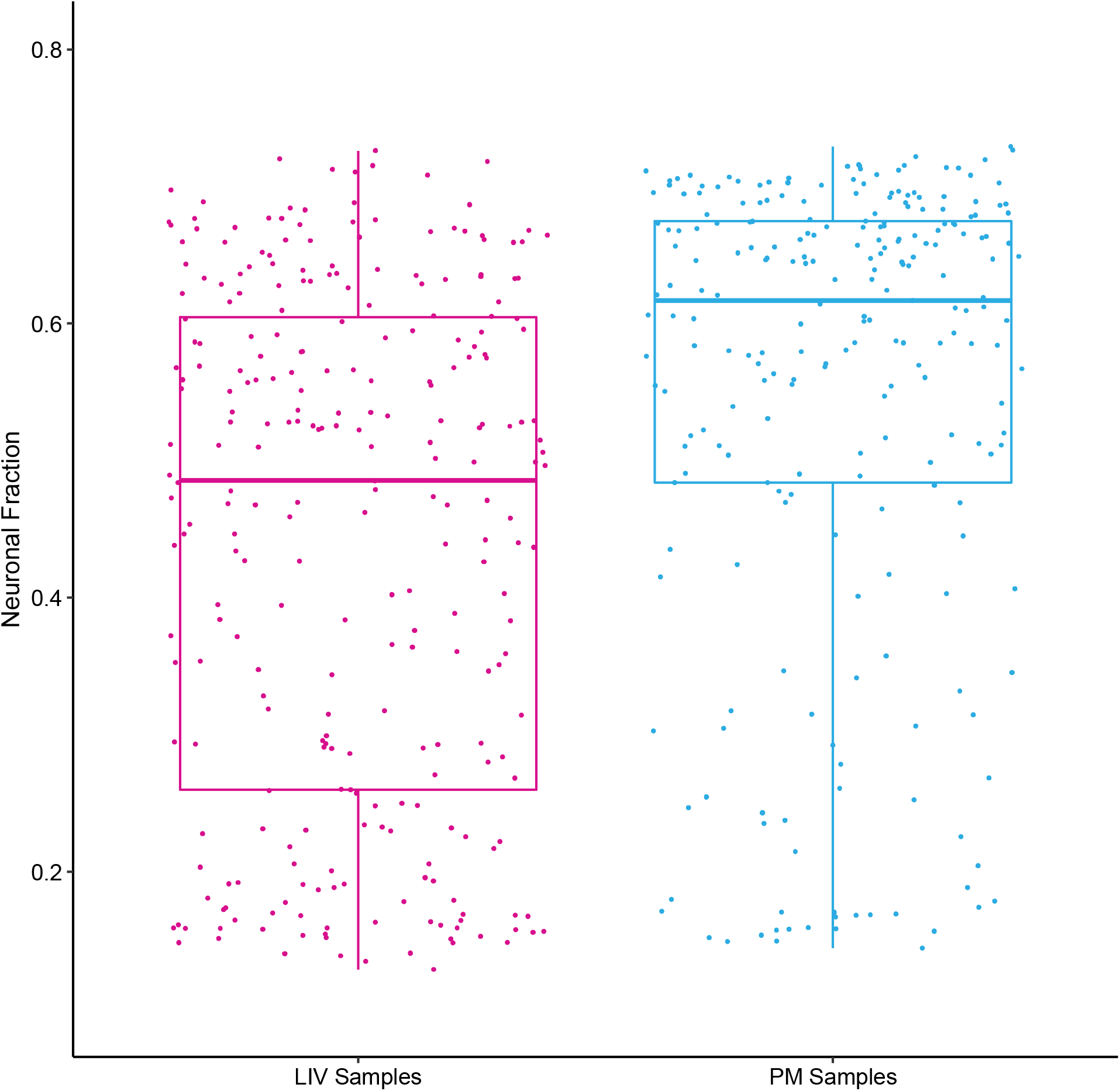
Neuronal cell fraction was calculated as the sum of the estimated glutamatergic and GABA-ergic neuronal fractions. The distributions of neuronal cell fractions are shown as boxplots for LIV samples (left) and PM samples (right). The upper and lower sides of each box represent the 75th and 25th percentiles of each distribution, respectively.

**Supplementary Figure 3:**
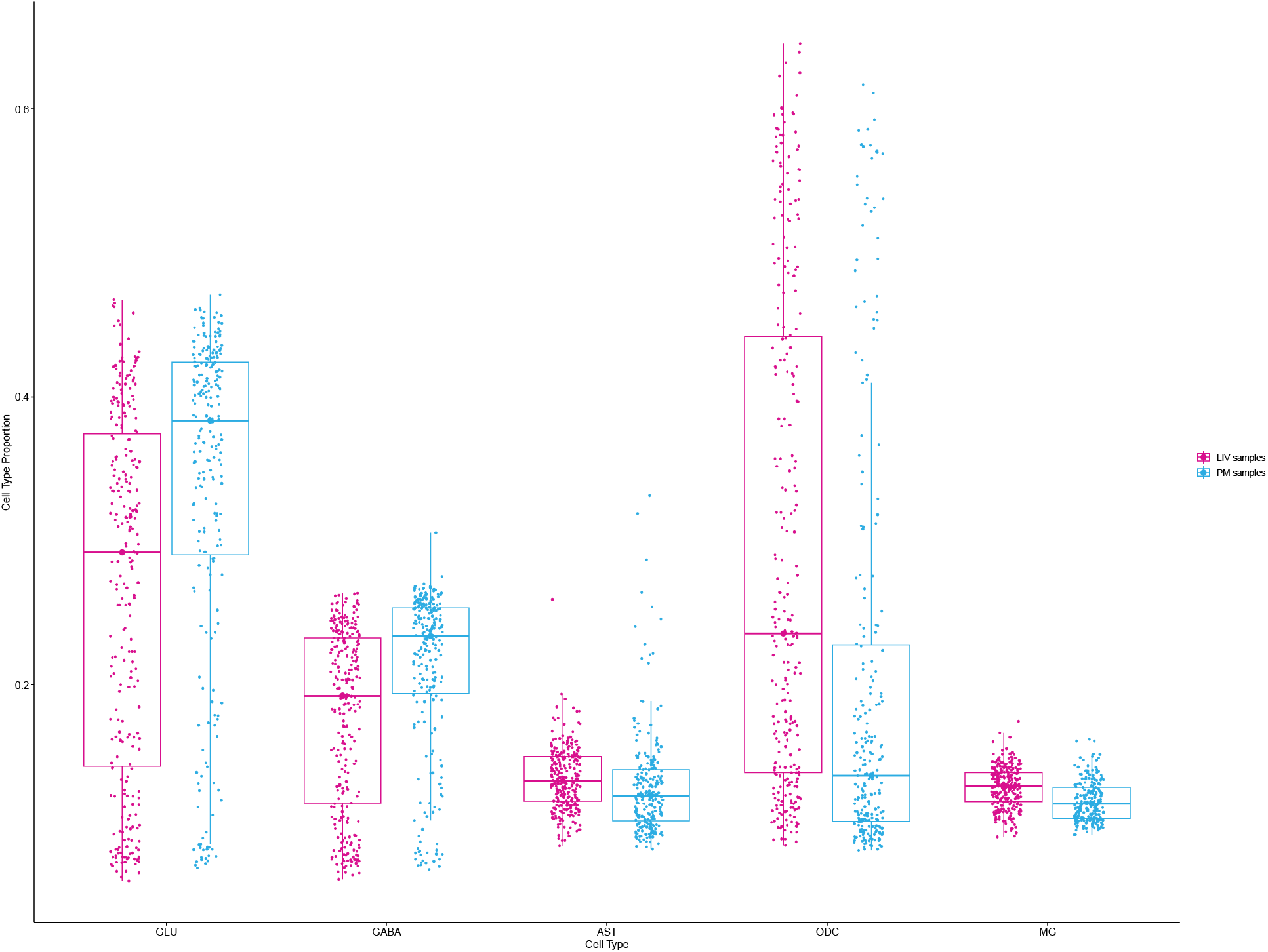
The distributions of estimated cell fractions are shown as boxplots for LIV samples (pink) and PM samples (blue). Distributions are shown for glutamatergic neurons (GLU), GABA-ergic neurons (GABA), astrocytes (AST), oligodendrocytes (ODC), and microglia (MG). The upper and lower sides of each box represent the 75th and 25th percentiles of each distribution, respectively.

**Supplementary Figure 4:**
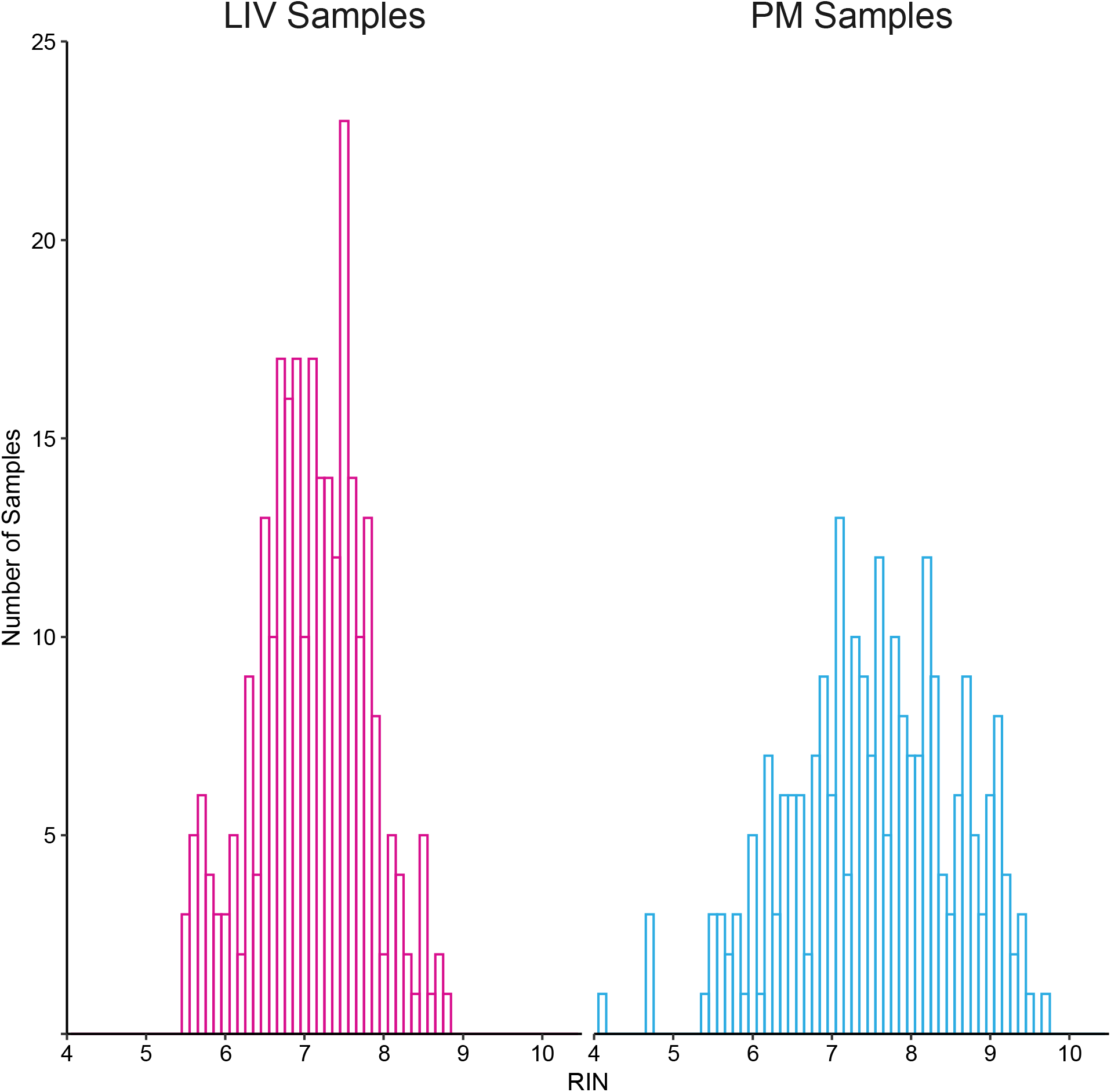
Histogram of RIN values for LIV samples and PM samples.

**Supplementary Figure 5:**
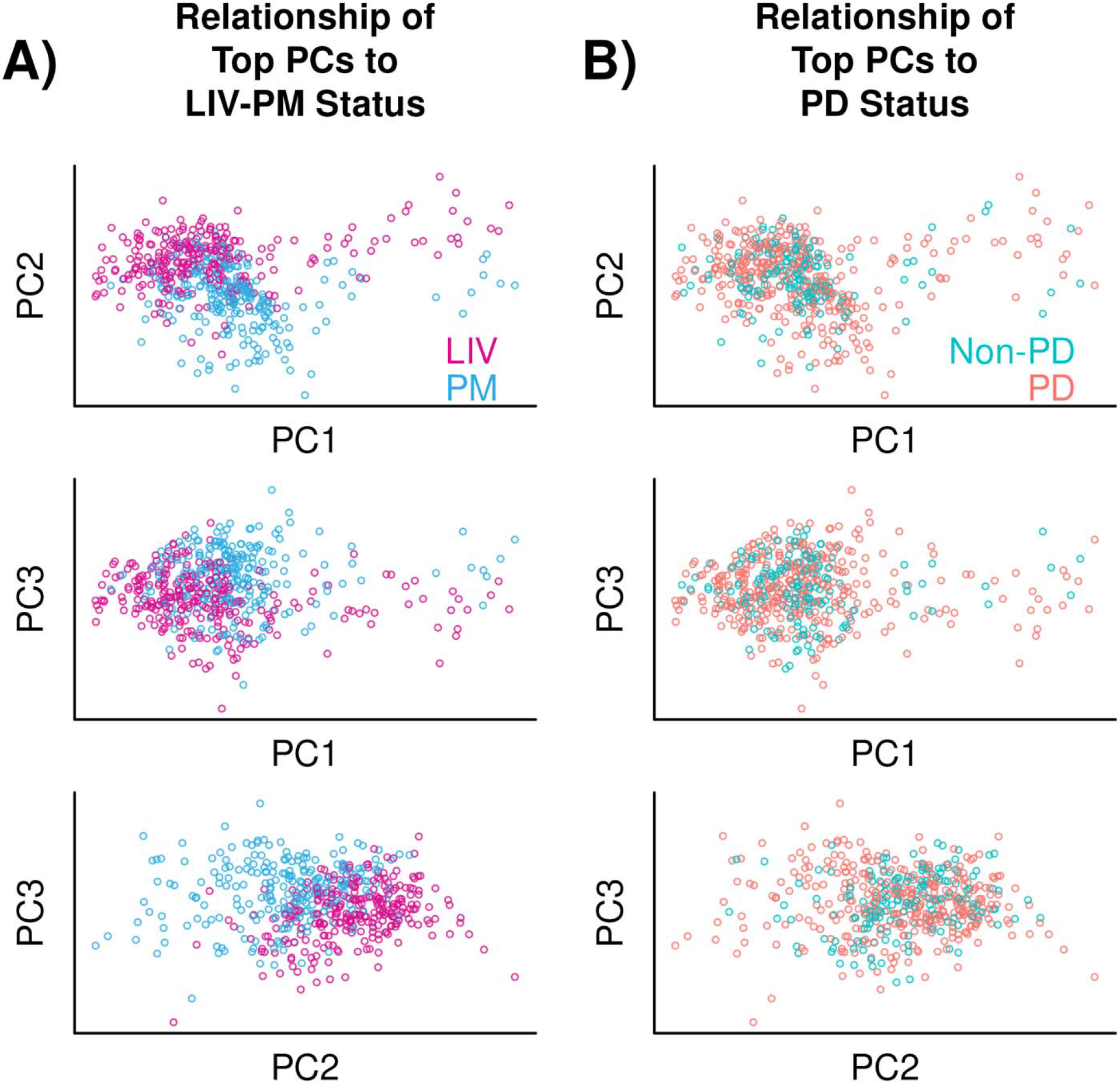
Principal components analysis. (A) Relationship of top PCs to LIV-PM status. Three scatter plots are presented showing the results of dimensionality reduction performed on the 21,635 genes expressed in the 518 samples in the full LBP cohort using principal components analysis. Each point is a sample, and colors differentiate LIV samples (pink) from PM samples (blue). The horizontal and vertical axes each represent the principal component (PC) indicated by the axis label. The top three PCs are shown, which collectively account for 27.20% of the variance in gene expression in the full LBP cohort (PC1 - 13.41%; PC2 - 7.41%; PC3 - 6.38%). (B) Relationship of top PCs to PD status. The same data plotted in A is shown except colors differentiate samples from individuals with PD (orange) from samples from individuals without PD (green).

**Supplementary Figure 6:**
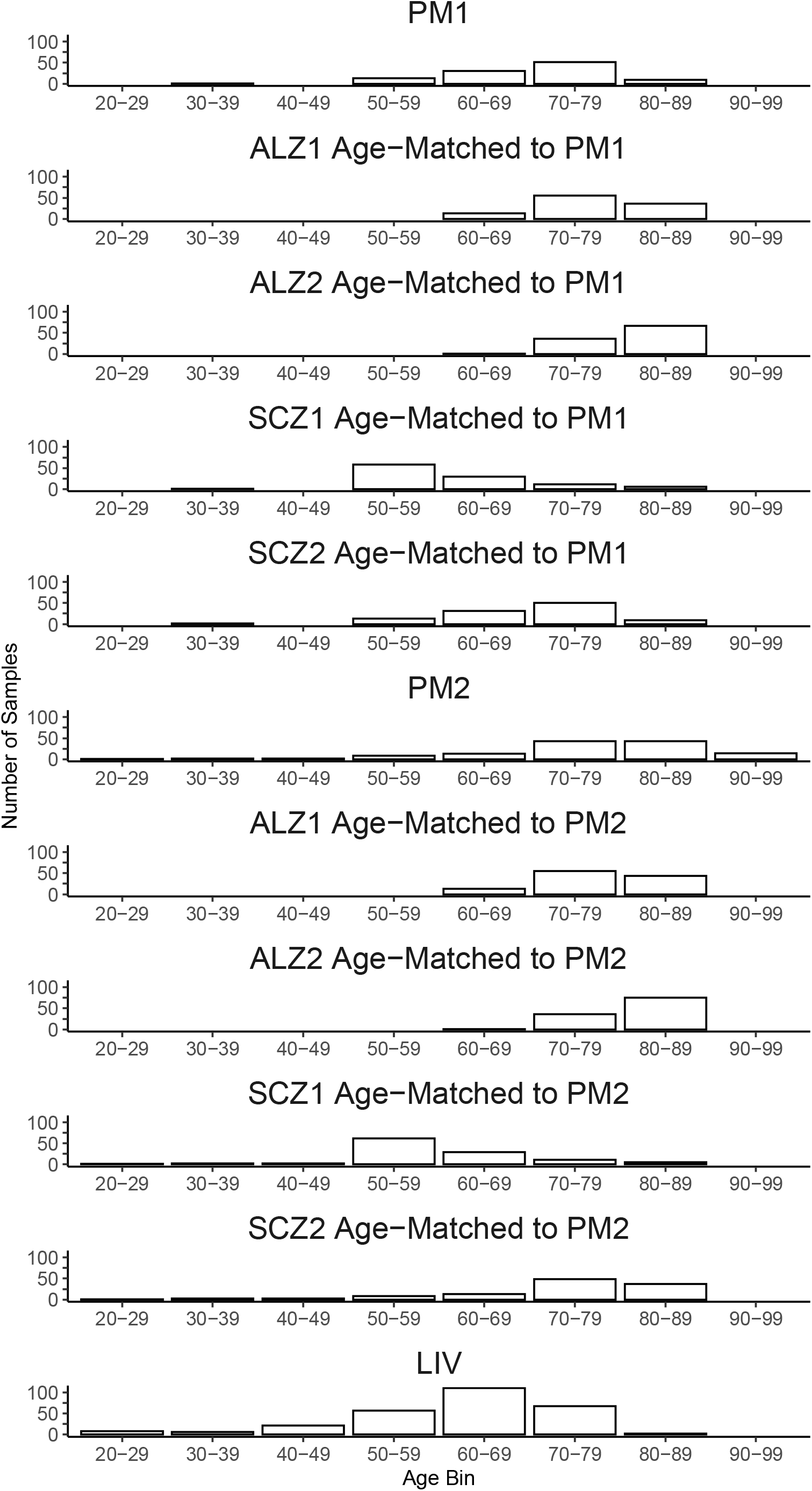
Propensity score matching was used to match the age distributions of the ALZ1, ALZ2, SCZ1, and SCZ2 datasets to the age distributions of the PM1 and PM2 datasets, respectively. For each of the ALZ1, ALZ2, SCZ1, and SCZ2 datasets, this resulted in two partially overlapping “age-matched” subsets – one composed of samples matched for age to the PM1 samples (“PM1 age-matched subset”; N = 104 samples) and one composed of samples matched for age to the PM2 samples (“PM2 age-matched subset”; N = 126 samples). Histograms of the age distributions are shown for the resulting PM1 age-matched subsets and PM2 age-matched subsets of the ALZ1, ALZ2, SCZ1, and SCZ2 datasets. Histograms are also shown for the age distributions of PM1 samples, PM2 samples, and LIV samples.

## Supplementary Table Captions

**Supplementary Table 1.**

The LIV-PM DE signature from the full LBP cohort (275 LIV samples and 243 PM samples)

**Supplementary Table 2.**

KEGG gene set enrichment results for LIV DEGs and PM DEGs in the LIV-PM DE signature from the full LBP cohort.

**Supplementary Table 3.**

Gene module assignments for the six co-expression networks constructed from subsets of the full LBP cohort.

**Supplementary Table 4.**

Results of testing five conserved network structures identified across LIV networks and PM networks for enrichment of LIV-PM DEGs and KEGG gene sets.

**Supplementary Table 5.**

DE signatures for ALZ, SCZ, PD, and Age.

**Supplementary Table 6.**

Results of testing for overlap of LIV-PM DEGs with ALZ, SCZ, PD, and Age DEGs.

**Supplementary Table 7.**

KEGG gene set enrichment results for the LIV PD structures enriched for PD GWAS genes.

## Methods

### Ethics statement

All human subjects research was carried out under STUDY-13-00415 of the Human Research Protection Program at the Icahn School of Medicine at Mount Sinai. Research participants in the living cohort provided informed consent for sample collection, genomic profiling, clinical data extraction from medical records, and public sharing of de-identified data.

### Living Brain Project cohort

PFC samples (N = 535; 289 LIV samples and 246 PM samples) were collected from 417 individuals (171 living, 246 postmortem).

#### LIV sample collection

LIV samples were collected from individuals undergoing the DBS electrode implantation procedure at the Icahn School of Medicine at Mount Sinai. Every English-speaking individual over 18 years old undergoing the procedure for any indication during the study period was eligible for enrollment. During the course of the procedure, a burr hole is drilled in the frontal bone in order to expose the cortical surface. To pass the electrode to the intended target, a cannula must be inserted into the exposed cortical surface. In order to prevent bleeding upon cannula insertion, the cortical surface is first “prepared” by sharply cutting the pial surface with a scalpel and cauterizing a circular region of the PFC. For the LBP, a simple modification was made to this standard clinical procedure: after the pial surface incision, a microdissector is used to obtain a small, circular biopsy of the PFC region that is typically destroyed by cauterization (***Figure 1B***). After the PFC biopsy is taken, the margins are cauterized such that the state of cortical surface preparation achieved is in effect equivalent to that achieved via the standard clinical procedure. With the margins cauterized, the neurosurgeon places the PFC biopsy on a dampened gauze pad, then hands the pad with the biopsy to research personnel outside the sterile field. While still in the operating room, the research personnel aliquot approximately one quarter of the biopsy into a tube of formalin and approximately three quarters of the biopsy into either a tube of RNAlater (Invitrogen, Waltham, MA) or a tube on dry ice. The aliquots are then immediately brought to the research laboratory where the smaller aliquot is embedded in paraffin and the larger aliquot is banked at −80°C.

#### Living cohort description

The majority of the LIV samples were obtained from individuals with PD, the most common indication for DBS. For these individuals, the diagnosis of PD was made by the treating neurologist and neurosurgeon based on established standards of care. Non-PD LIV samples were obtained from individuals undergoing the DBS procedure for indications other than PD. The number of living participants for each DBS indication is provided in ***Figure 1C***. Unilateral biopsies were obtained from 53 living LBP participants (40 from the left hemisphere and 13 from the right hemisphere) and bilateral biopsies were obtained from 118 living LBP participants. Bilateral biopsies were obtained one month apart (left hemisphere followed by right hemisphere) due to the staged approach to DBS electrode implantation that is the standard of care at the Icahn School of Medicine at Mount Sinai. Blood was obtained by venipuncture concurrently with the PFC biopsy in most instances.

#### Postmortem cohort description

Frozen postmortem PFC samples were obtained from three separate brain banks: Harvard Brain and Tissue Resource Center (“PM1” in text, figures, and tables; N = 104), the New York Brain Bank at Columbia University (“PM2”; N = 129), and the University of Miami Brain Endowment Bank (“PM3”; N = 13). PM samples were matched to LIV samples for age and sex to the greatest extent possible (***Figure 1C***). A portion of each frozen PM sample was formalin-fixed and embedded in paraffin.

PM1 and PM3 samples were acquired through National Institutes of Health NeuroBioBank (NIH NBB) request #543. Sample processing following autopsies at NIH NBB repositories follows standard protocols that have been described elsewhere^37,38^. In brief, one hemisphere (or selected bilateral blocks) is dissected into coronal sections approximately 0.5-1.0 centimeters (cm) thick, flash-frozen (-150°C), and stored at −80°C. Informed consent is obtained from donors who register before death and/or from donors’ next-of-kin. All policies and procedures are reviewed and approved by the respective repository’s institutional review board and any additional institutional oversight committees as necessary and appropriate. Diagnoses are made based on information obtained from medical records, questionnaires, and/or interviews with individuals with knowledge of the donor, and from neuropathological evaluation.

PM2 samples were acquired through New York Brain Bank at Columbia University request #1962. New York Brain Bank at Columbia University processes the brains of donors with age-related neurodegenerative diseases, and donors without neurologic or psychiatric impairments^39^. The donors were evaluated at Columbia University healthcare facilities. The protocol used for sample processing has been described in detail elsewhere^40^. In brief, one standardized series of 18 blocks (ranging in size between 0.3 × 0.5 × 1.0 cm and 0.8 × 2.5 × 3.0 cm) from 18 standardized anatomic sites is harvested from each half of the brain and immediately deep-frozen using liquid nitrogen vapor (-160 to −180°C). Gross neuropathologic examination is performed during processing. Diagnoses are made by a consensus team following discussion of all available clinical and pathological information.

#### Anesthesia in living cohort

Up to three anesthetics were administered to living participants for DBS surgery: dexmedetomidine, fentanyl, and/or propofol. For 281 of the 289 LIV samples, information regarding the anesthetics administered during the corresponding DBS surgery was extracted from the electronic medical record. For each anesthetic, the total dose given to the participant was calculated as the dose administered by infusion plus the dose administered by bolus.

#### PD symptom severity in living cohort

To assess PD symptom severity, the Unified Parkinson’s Disease Rating Scale (UPDRS) Part III Motor Examination (“UPDRS-III”) was administered to living participants with PD as part of routine clinical care for DBS. UPDRS-III scores were able to be extracted from the electronic medical record for 107 of the 132 living individuals with PD. The UPDRS-III was administered when individuals were off dopamine replacement therapy (i.e., levodopa) prior to the first DBS surgery (median time between UPDRS-III administration and first DBS surgery = 60 days). UPDRS-III scores less than 40 were defined as low and UPDRS-III scores >= 40 were defined as high. The threshold for defining high and low UPDRS-III scores was chosen based on manual inspection of the distribution of the 107 UPDRS-III scores.

#### Dopamine replacement therapy

Levodopa dosage information was able to be extracted from the electronic medical record for 130 of the 132 living individuals with PD. Levodopa dosage information was obtained either at the time of UPDRS-III administration or at another time point in the weeks prior to the participant’s first DBS surgery. If the participant was receiving more than one levodopa medication (e.g., generic levodopa and name-brand levodopa), the total levodopa dose was calculated by summing the doses of all levodopa medications. Levodopa dose less than 900 milligrams (mg) was defined as low and levodopa dose greater than 900 mg was defined as high. The threshold for defining high and low levodopa doses was chosen based on the median of the distribution of the 130 levodopa doses.

#### Lewy body staining

Staining for Lewy bodies was performed on subsets of the LIV samples (N = 102) and PM samples (N = 93) at the Icahn School of Medicine at Mount Sinai. Formalin-fixed paraffin-embedded tissue slices approximately 5 micrometers (μm) thick were baked at 70 – 80°C on charged slides for an average of 15 minutes. Chromogenic immunohistochemistry was performed on the BOND Rx Automated Research Stainer (Leica Biosystems, Wetzlar, Germany), as follows:

1. Heat-induced epitope retrieval was performed at a pH of 9 on the slides for 20 minutes
2. Slides were then incubated in the primary antibody, mouse anti-alpha-synuclein (1:6000 antibody to diluent, LB509, MABN824MI, MilliporeSigma) for 30 minutes
3. The slides were then incubated for 20 minutes with the BOND Polymer Refine Detection kit (DS9800, Leica Biosystems, Wetzlar, Germany), which uses 3,3’-Diaminobenzidine (DAB) chromogen and hematoxylin counterstain to visualize the primary antibody in the tissue section.

Whole slide images were scanned at 40x magnification using the Versa 8 Scanner (Leica Biosystems, Wetzlar, Germany).

#### Lewy body scoring

A laboratory technician blinded to the clinical histories of living participants and postmortem donors reviewed each whole slide image for the presence and density of Lewy bodies. Whole slide images were given a score of 0 (no evidence of Lewy neurites or bodies), 1-(some Lewy neurites, no Lewy bodies), 1 (1 Lewy body), 1+ (2 Lewy bodies), 2-(3-6 Lewy bodies), 2 (7-9 Lewy bodies), 2+ (10-12 Lewy bodies), 3-(13-15 Lewy bodies), 3 (16-18 Lewy bodies), or 3+ (>= 19 Lewy bodies). Scoring criteria were relative to tissue sections of size 1500 μm x 1500 μm and were adjusted for tissue sections of larger or smaller size. For DE analyses, scores were grouped as follows: “No Lewy bodies” (score of 0), “Low Lewy body density” (score of 1-), “Medium Lewy body density” (scores of 1 and 1+), “High Lewy body density” (scores of 2- and above). Scores were grouped in this manner to create sample sizes sufficiently powered for DE analysis.

### Batch assignments for LBP sample processing

To minimize batch effects, all LIV samples (N = 289), PM samples (N = 246), and blood samples obtained from living LBP participants (N = 244; used in this report only to identify mislabeled PFC samples) were processed together for RNA sequencing at the Icahn School of Medicine at Mount Sinai. Samples were assigned to batches for RNA extraction, cDNA library preparation, and RNA sequencing using a randomization algorithm that minimized correlations between batch assignments and LIV-PM status. For extraction, batches of size 12 were constructed via the following procedure: (1) a list of LIV samples and a list of PM samples to be randomized is defined, (2) the smaller of these two lists is iterated through, (3) at each iteration a pair is created of one randomly selected LIV sample and one randomly selected PM sample, (4) sample pairs are randomly combined into batches of size 12. RNA extraction of blood samples was performed separately from RNA extraction of PFC samples due to different extraction kits required. For cDNA library preparation, a similar procedure was performed but batches of size 32 were created and instead of creating pairs comprised of one LIV sample and one PM sample, trios were created comprised of one LIV sample, one PM sample, and one blood sample. Batches of size 96 were constructed for RNA sequencing by randomly grouping three cDNA library preparation batches together. The creation of pairs and trios in the manner described ensured there was an even distribution of LIV samples and PM samples across batches for all of the sample processing steps. For both the RNA extraction and cDNA library preparation steps, 1,000,000 permutations of the batch assignment procedure were performed and for each permutation the correlation between batch assignment and several variables of interest (e.g., LIV-PM status) was determined using the canCorPairs() function of the variancePartition R package (v1.20.0)^41^. Higher correlation between batch and a variable of interest implies stronger batch effects, so batch assignment permutations were selected that minimized correlation over these variables, with priority given to minimizing the correlation between batch assignments and LIV-PM status. Effort was made to adhere to the selected batch assignments at every processing step, but for a minority of batches the actual composition deviated slightly from the assignments made by the algorithm.

### Sample preparation and RNA extraction

Approximately 5-10 mg of each LIV sample and PM sample were used for RNA extraction. As a size reference for aliquoting the 535 specimens for RNA extraction, a piece of PM brain tissue was cut, weighed, and placed in a clear Eppendorf tube on dry ice. The reference was used throughout the whole extraction process to eliminate the need for weighing individual samples, which would result in thawing and RNA degradation. To maintain all materials at a uniform temperature, a cryostat was set to −20°C and tools for aliquoting (e.g., blades, petri dishes, forceps) were placed in the chamber, preventing brain tissue from thawing while handling. All equipment that could come into contact with tissue (e.g., cryostat platform, gloves) was treated with RNase Zap (Invitrogen, Waltham, MA; Catalog Number AM9780). Each sample was cut to the reference size in the cryostat and placed in an empty pre-labeled Eppendorf tube on dry ice. After a batch of 12 samples had been aliquoted, samples were removed from dry ice and immediately pipetted into 500 microliters (μL) of TRIzol (Invitrogen, Waltham, MA; Catalog Number 15596026) in the Eppendorf tube. Samples were then homogenized in TRIzol using the Tissue Ruptor (Qiagen, Germany), pulsing for one second on the highest speed three times or until the sample was completely homogenized (i.e., no tissue was visible). RNA extraction for 528 of the 535 LBP specimens was performed with the RNeasy Kit (Qiagen, Hilden, Germany), mostly according to manufacturer instructions. For six samples, extraction was performed using the Illustra TriplePrep Kit (Cytiva Life Sciences, Marlborough, MA; Catalog Number 28-9425-44) and for one sample the extraction kit used was not recorded (these seven samples were not part of the randomization procedure for extraction batch assignment). The RNA isolate was then stored at −80°C. RNA fragment sizes were assessed using the 2100 BioAnalyzer System (Agilent, Santa Clara, USA) and only specimens with an RNA integrity number (RIN) greater than 4.0 were sent for sequencing.

### RNA sequencing

Preparation of cDNA libraries and RNA sequencing were performed at Sema4 (Stamford, CT). Libraries of cDNA were prepared using the TruSeq Stranded Total RNA with Ribo-Zero Globin Kit (Illumina, San Diego, CA; Catalog Number 20020613). RNA sequencing was performed on the NovaSeq 6000 System (Illumina, San Diego, CA). For each sequencing batch, two S4 flow cells were used to achieve a targeted depth of 100 million 100-base-pair paired-end reads per sample library. The 96 barcoded sample libraries in each batch were pooled and loaded into each of the four lanes of both flow cells. Sequencing was performed in two waves. In the first wave, samples were sequenced to a depth of 50 million paired-end reads. In the second wave over a year later, the remaining samples were sequenced to a depth of 100 million, and an additional 50 million reads were generated on the samples from the first wave.

### RNA sequencing data alignment, quantification, and normalization

The RAPiD pipeline^42^ was used for processing RNA sequencing data as previously described^3,11,43,44^. Base calls made from clusters within the S4 flow cell were organized into sequencing reads and stored in FASTQ files using Illumina’s bcl2fastq software^45^. These unaligned reads were then aligned to the GRCh38 primary assembly^46^ with Gencode gene annotation (v30)^46^ using STAR (v2.7.2a)^47^. The following non-default STAR parameters were used (defaults were used for all other parameters): --sjdbScore 1, --outSAMstrandField intronMotif, --outFilterMismatchNoverReadLmax 0.04, --outReadsUnmapped Fastx, --outSAMunmapped Within, --outSAMtype BAM Unsorted, --outSAMattributes NH HI AS NM MD, --quantMode TranscriptomeSAM. For a given sample, all FASTQs (i.e., the FASTQs from both flow cells the sample was sequenced on) were aligned together. Aligned reads were sorted using samtools (v1.13)^48^ and duplicate reads were marked using Picard Tools (v2.20.1)^49^. Gene expression quantification was done with featureCounts (Linux subread package v1.6.3)^50^ using primary alignments only, strandedness option set to 2, feature type set to exon, attribute type set to gene_id, and count all overlapping features set to true. Lowly expressed genes were defined as genes with less than 1 count per million in at least 10% of samples and removed from analysis. The raw counts of expressed genes were normalized using the voomWithDreamWeights() function of the dream software^51^ within the variancePartition R package because two samples per individual were available for some individuals. Since the data from the discovery wave, which was used to identify the LIV-PM DE signature in the discovery wave cohort, consisted of one sample per individual, the raw counts of expressed genes were normalized using the voom function of the limma R package (v3.46.0)^52^.

### RNA sequencing data cell type composition

Cell type deconvolution was performed on the gene expression counts data prior to normalization using the dtangle R package (v2.0.9)^53^. Single-cell RNA sequencing data from postmortem human PFC cells was used as the cell type reference^17^ – specifically data from astrocytes (N = 737 cells), GABA-ergic neurons (N = 2,337), glutamatergic neurons (N = 4,771), microglia (N = 187), and oligodendrocytes (N = 2,191). The neuronal cell fraction estimate was calculated by adding the glutamatergic and GABA-ergic neuronal cell fraction estimates.

### RNA sequencing data confounder identification

Technical metrics characterizing the RNA sequencing data were generated for quality control purposes using STAR, featureCounts, Picard Tools, and fastqc (v0.11.8)^54^ implemented via the RAPiD pipeline. These metrics were merged into a table of covariates that included variables characterizing individuals (e.g., age, sex) and samples (e.g., batch assignments, RIN values). Confounders explaining the variance in gene expression between samples were then identified to be included as covariates in downstream analyses through an adaptation of an iterative procedure established in previous RNA sequencing studies^12^. First, LIV-PM status, RIN, individual ID, and sex were fit to a linear mixed model with individual ID and sex as random effects using the dream() function of the variancePartition R package, and residuals were calculated with the residuals() function of the base stats R package. For processing the data from the discovery wave used in the LIV-PM DE analysis of the discovery wave cohort, the lmFit() function of the limma R package was used for this step and individual ID was not added as there was only one sample per individual. RIN, individual ID, and sex were accounted for because these variables are known to influence gene expression. Accounting for the effects of LIV-PM status on the variance in gene expression at this step allowed the effects of other covariates on the variance to subsequently be observed. Next, principal component analysis (PCA) was performed on the covariance of the residual gene expression matrix using the prcomp() function of the base stats R package and the canonical correlation between the variables in the covariate table and principal components (PCs) 1-5 of the residual expression data was calculated using the canCorPairs() function of the variancePartition R package. Scatterplots of PCs (e.g., PC1 vs PC2) were generated and colored for each covariate as a visual aid in assessing the strength of the relationships between covariates and expression data. Canonical correlations and visual aids were reviewed and one covariate – for which the correlation with LIV-PM status was minimized and the correlation with PC1 was maximized – was selected to be included in downstream models. Technical covariates with high correlations to LIV-PM status were purposefully not selected during the latter step because the even distribution of LIV samples and PM samples across batches (achieved via the randomization procedure) made it more likely that these correlations were driven by true biological differences between LIV samples and PM samples rather than technical confounders introduced during experimental procedures. The selected covariate was added to the linear model and the process was repeated until no additional covariates could be identified through this procedure as affecting the variance in gene expression. Additional covariates affecting variance in gene expression in ways that could not be identified by this procedure were identified using a network approach. A co-expression network was built from the residual gene expression matrix (accounting for the set of covariates found via the process just described) using the WGCNA R package (v1.69)^55^, and correlations between covariates and network module eigengenes were calculated. The correlations of covariates with (1) module eigengenes, (2) other covariates, and (3) LIV-PM status were considered, and an iterative process was used to select for addition to the model the fewest number of covariates with minimal association to LIV-PM status and maximal association to module eigengenes.

### RNA sequencing data sample filtering

Samples were removed either for being outliers in the expression data or for being identified as mislabeled without a definitive way to correct their labels. Outliers were defined as samples falling more than three standard deviations away from the centroid of PC1 and PC2 of the residual gene expression matrix after accounting for covariates selected using the above procedures. To identify sample mislabeling events, identity concordance between samples was performed using genetic variants called from the PFC RNA sequencing data as well as from blood RNA sequencing data and microarray-based genotyping data generated from the same individuals for other ongoing studies. More than one data source was available for identity concordance for all but one individual. Variants were called from RNA sequencing data following Genome Analysis Toolkit (GATK) best practices^56^. The approach used to determine if two variant call sets were from the same individual differed depending on the sources of the call sets in the comparison (i.e., RNA sequencing data to RNA sequencing data, microarray genotyping data to microarray genotyping data, or RNA sequencing data to microarray genotyping data). For comparisons of two call sets from RNA sequencing data or two call sets from microarray genotyping data, gtcheck from the bcftools software package (v1.9) was used to calculate the percentage of sites concordant between the call sets. For comparisons of RNA sequencing data call sets to microarray genotyping data call sets, genotyping matrices were read into R, discrepancies between allele code fields were corrected, sites covered in only one call set were removed, and Pearson’s correlations of genotype similarity were calculated for every pair of call sets. Regardless of the approach used to calculate concordance between call sets, a correlation threshold for deciding whether two samples came from the same individual was determined manually by assessing the distribution of similarity metrics. Mismatches were defined as (1) instances where two samples expected to be from the same individual were genetically discordant and (2) instances where two samples not expected to be from the same individual were genetically concordant. All mismatches identified by the thresholding procedure were further examined to confirm a true mismatch (i.e., using all RNA sequencing data and microarray genotyping data call sets from the individuals in the mismatch). Following sample filtering, 518 samples were retained for analysis (“the full LBP cohort”).

### Differential expression analyses in LIV and PM samples

A total of 66 DE analyses were performed on LIV samples and PM samples for this report, including DE of (1) LIV-PM status, (2) age, (3) the interaction between LIV-PM status and age, and (4) PD status. For most DE analyses performed on the LIV and PM samples, DE was run on a normalized count matrix using the dream() function in the variancePartition R package and one of the formulas indicated below. The two exceptions were (1) the LIV-PM DE analysis performed in the discovery wave cohort, which was performed using the lmFit() function in the limma R package, and (2) the LIV-PM DE analysis performed using the DESeq() function in the DESeq2 R package to assess the concordance of LIV-PM DE signatures generated using different DE methods. For all DE analyses performed on LIV and PM samples, multiple test correction using an FDR of 5% was computed by the DE software package.

The 66 DE analyses performed in LIV samples and PM samples in this report are presented in the list that follows. For each DE analysis on the list, the formula used for DE is indicated in square brackets. All formulas are provided below the list. The details of these DE analyses (including the nomenclature of these DE analyses) are provided in their respective sections in the Results and Supplementary Information.

List of 66 DE analyses performed:

1. LIV-PM DE to identify the primary LIV-PM DE signature (Figure 2A, Supplementary Table 1) [Formula 1]
2. LIV-PM DE in the discovery wave cohort (Figure 2B) [Formula 2]
3. LIV-PM DE in the replication wave cohort (Figure 2B) [Formula 1]
4. LIV-PM DE in LIV PD samples and PM PD samples only (Figure 2B) [Formula 1]
5. LIV-PM DE in LIV non-PD samples and PM non-PD samples only (Figure 2B) [Formula 1]
6. LIV-PM DE in PM1 samples and half of LIV samples (Figure 2B) [Formula 1]
7. LIV-PM DE in PM2 samples and half of LIV samples (Figure 2B) [Formula 1]
8. LIV-PM:*LowPMI1* DE (Figure 2B) [Formula 1]
9. LIV-PM:*HighPMI* DE (Figure 2B) [Formula 1]
10. LIV-PM:*LowPMI2* DE (Figure 2B) [Formula 1]
11. LIV-PM:*HighPMI2* DE (Figure 2B) [Formula 1]
12. PD1 DE (i.e., PD DE in PM1) (Figure 4A, Supplementary Table 5) [Formula 4]
13. PD2 DE (i.e., PD DE in PM2) (Figure 4A, Supplementary Table 5) [Formula 4]
14. LIV Age DE (Figure 4A, Supplementary Table 5) [Formula 5]
15. PM1 Age DE (Figure 4A, Supplementary Table 5) [Formula 5]
16. PM2 Age DE (Figure 4A, Supplementary Table 5) [Formula 5]
17. Age/LIV-PM status interaction DE [Formula 6]
18. Lewy body pathology: LIV*:No*-PM*:Low* DE [Formula 1]
19. Lewy body pathology: LIV*:No*-PM:*Med* DE [Formula 1]
20. Lewy body pathology: LIV*:No*-PM*:High* DE [Formula 1]
21. Lewy body pathology: LIV*:Low*-PM*:Low* DE [Formula 1]
22. Lewy body pathology: LIV*:Low-*PM*:Med* DE [Formula 1]
23. Lewy body pathology: LIV*:Low*-PM*:High* DE [Formula 1]
24. Lewy body pathology: LIV*:No*-LIV:*Low* DE [Formula 1]
25. Lewy body pathology: PM*:Low*-PM:*Med* DE [Formula 1]
26. Lewy body pathology: PM*:Low*-PM:*High* DE [Formula 1]
27. Lewy body pathology: PM*:Med*-PM:*High* DE [Formula 1]
28. Anesthesia: LIV*:PropofolQ1*-PM DE [Formula 1]
29. Anesthesia: LIV*:PropofolQ2*-PM DE [Formula 1]
30. Anesthesia: LIV*:PropofolQ3*-PM DE [Formula 1]
31. Anesthesia: LIV*:PropofolQ4*-PM DE [Formula 1]
32. Anesthesia: LIV*:DexmedetomidineQ1*-PM DE [Formula 1]
33. Anesthesia: LIV*:DexmedetomidineQ2*-PM DE [Formula 1]
34. Anesthesia: LIV*:DexmedetomidineQ3*-PM DE [Formula 1]
35. Anesthesia: LIV*:DexmedetomidineQ4*-PM DE [Formula 1]
36. Anesthesia: LIV*:FentanylQ1*-PM DE [Formula 1]
37. Anesthesia: LIV*:FentanylQ2*-PM DE [Formula 1]
38. Anesthesia: LIV*:FentanylQ3*-PM DE [Formula 1]
39. Anesthesia: LIV*:FentanylQ4*-PM DE [Formula 1]
40. LIV:*RNAlater*-PM DE [Formula 1]
41. LIV:*DryIce*-PM DE [Formula 1]
42. LIV:*lowUPDRS*-PM DE [Formula 1]
43. LIV:*highUPDRS*-PM DE [Formula 1]
44. LIV:*lowDOPA*-PM DE [Formula 1]
45. LIV:*highDOPA*-PM DE [Formula 1]
46. LIV-PM DE using “the 50% definition” of expressed genes [Formula 1]
47. LIV-PM DE using “the 75% definition” of expressed genes [Formula 1]
48. LIV-PM DE with DESeq2 [Formula 3]
49. LIV:*lowRIN*-PM:*lowRIN* DE [Formula 1]
50. LIV:*highRIN*-PM:*highRIN* DE [Formula 1]
51. LIV:*lowAge*-PM:*lowAge* DE [Formula 1]
52. LIV:*highAge*-PM:*highAge* DE [Formula 1]
53. LIV:*lowAge*-PM:*highAge* DE [Formula 1]
54. LIV:*highAge*-PM:*lowAge* DE [Formula 1]
55. LIV-PM DE with ODC fraction as the *Cell Type Fraction* in the formula [Formula 7]
56. LIV-PM DE with AST fraction as the *Cell Type Fraction* in the formula [Formula 7]
57. LIV-PM DE with MG fraction as the *Cell Type Fraction* in the formula [Formula 7]
58. LIV-PM DE with neuronal fraction as *Cell Type Fraction 1* and ODC fraction as *Cell Type Fraction 2* in the formula [Formula 8]
59. LIV-PM DE with neuronal fraction as *Cell Type Fraction 1* and AST as *Cell Type Fraction 2* in the formula [Formula 8]
60. LIV-PM DE with neuronal fraction as *Cell Type Fraction 1* and MG as *Cell Type Fraction 2* in the formula [Formula 8]
61. LIV-PM DE with ODC fraction as *Cell Type Fraction 1* and AST as *Cell Type Fraction 2* in the formula [Formula 8]
62. LIV-PM DE with ODC fraction as *Cell Type Fraction 1* and MG fraction as *Cell Type Fraction 2* in the formula [Formula 8]
63. LIV-PM DE with AST fraction as *Cell Type Fraction 1* and MG as *Cell Type Fraction 2* in the formula [Formula 8]
64. LIV-PM DE with neuronal fraction as *Cell Type Fraction 1*, ODC fraction as *Cell Type Fraction 2*, and MG fraction as *Cell Type Fraction 3* in the formula [Formula 9]
65. LIV-PM DE with neuronal fraction as *Cell Type Fraction 1*, AST fraction as *Cell Type Fraction 2*, and MG fraction as *Cell Type Fraction 3* in the formula [Formula 9]
66. LIV-PM DE with ODC fraction as *Cell Type Fraction 1*, AST fraction as *Cell Type Fraction 2*, and MG fraction as *Cell Type Fraction 3* in the formula [Formula 9]

#### Formula 1 (linear mixed model with fixed effects and random effects for individual ID, depletion batch, and sex)

*Gene Expression ∼ LIV-PM Status + Individual ID + Sex + RIN + Neuronal Fraction + Picard RNASeqMetrics Median 3’ Bias + Picard RNASeqMetrics PCT mRNA Bases + Depletion Batch + Picard InsertSizeMetrics Median Insert Size + Picard AlignmentSummaryMetrics Strand Balance (First of Pair) + Residuals*

#### Formula 2 (linear model with fixed effects only)

*Gene Expression ∼ LIV-PM Status + Sex + RIN + Neuronal Fraction + Residuals*

#### Formula 3 (linear model with fixed effects only)

*Gene Expression ∼ LIV-PM Status + Sex + RIN + Neuronal Fraction + Picard RNASeqMetrics Median 3’ Bias + Picard RNASeqMetrics PCT mRNA Bases + Depletion Batch + Picard InsertSizeMetrics Median Insert Size + Picard AlignmentSummaryMetrics Strand Balance (First of Pair) + Residuals*

#### Formula 4 (linear model with fixed effects only)

*Gene Expression ∼ PD Status + Sex + RIN + Neuronal Fraction + Picard RNASeqMetrics Median 3’ Bias + Picard RNASeqMetrics PCT mRNA Bases + Depletion Batch + Picard InsertSizeMetrics Median Insert Size + Picard AlignmentSummaryMetrics Strand Balance (First of Pair) + Residuals*

#### Formula 5 (linear mixed model with fixed effects and random effects for individual ID, depletion batch, sex, and PD status)

*Gene Expression ∼ Age + PD Status + Sex + RIN + Neuronal Fraction + Picard RNASeqMetrics Median 3’ Bias + Picard RNASeqMetrics PCT mRNA Bases + Individual ID + Depletion Batch + Picard InsertSizeMetrics Median Insert Size + Picard AlignmentSummaryMetrics Strand Balance (First of Pair) + Residuals*

#### Formula 6 (linear mixed model with fixed effects and random effects for individual ID, depletion batch, sex, and PD status)

*Gene Expression ∼ Age + LIV-PM Status + Age*LIV-PM Status + PD Status + Sex + RIN + Neuronal Fraction + Picard RNASeqMetrics Median 3’ Bias + Picard RNASeqMetrics PCT mRNA Bases + Individual ID + Depletion Batch + Picard InsertSizeMetrics Median Insert Size + Picard AlignmentSummaryMetrics Strand Balance (First of Pair) + Residuals*

#### Formula 7 (linear mixed model with fixed effects and random effects for individual ID, depletion batch, and sex)

*Gene Expression ∼ LIV-PM Status + Individual ID + Sex + RIN + Cell Type Fraction + Picard RNASeqMetrics Median 3’ Bias + Picard RNASeqMetrics PCT mRNA Bases + Depletion Batch + Picard InsertSizeMetrics Median Insert Size + Picard AlignmentSummaryMetrics Strand Balance (First of Pair) + Residuals*

#### Formula 8 (linear mixed model with fixed effects and random effects for individual ID, depletion batch, and sex)

*Gene Expression ∼ LIV-PM Status + Individual ID + Sex + RIN + Cell Type Fraction 1 + Cell Type Fraction 2 + Picard RNASeqMetrics Median 3’ Bias + Picard RNASeqMetrics PCT mRNA Bases + Depletion Batch + Picard InsertSizeMetrics Median Insert Size + Picard AlignmentSummaryMetrics Strand Balance (First of Pair) + Residuals*

#### Formula 9 (linear mixed model with fixed effects and random effects for individual ID, depletion batch, and sex)

*Gene Expression ∼ LIV-PM Status + Individual ID + Sex + RIN + Cell Type Fraction 1 + Cell Type Fraction 2 + Cell Type Fraction 3 + Picard RNASeqMetrics Median 3’ Bias + Picard RNASeqMetrics PCT mRNA Bases + Depletion Batch + Picard InsertSizeMetrics Median Insert Size + Picard AlignmentSummaryMetrics Strand Balance (First of Pair) + Residuals*

### Principal components analysis

The PCA presented in ***Supplementary Figure 5*** was performed using the prcomp() function of the base stats R package. Residual gene expression data that was input to the prcomp() function was calculated from the full LBP cohort using the following formula:

#### Formula 10 (linear model with fixed effects and random effects for individual ID, depletion batch, and sex)

*Gene Expression ∼ Individual ID + Sex + RIN + Neuronal Fraction + Picard RNASeqMetrics Median 3’ Bias + Picard RNASeqMetrics PCT mRNA Bases + Depletion Batch + Picard InsertSizeMetrics Median Insert Size + Picard AlignmentSummaryMetrics Strand Balance (First of Pair) + Residuals*

### Biological annotation of LIV-PM DE signature

Housekeeping genes were obtained from E. Eisenberg and E.Y. Levanon (2013)^19^. All gene set enrichment analyses were performed using the Kyoto Encyclopedia of Genes and Genomes (KEGG) database^20^. The htext-formatted hsa00001 KEGG database was downloaded on October 4th, 2021, via the KEGG web server and parsed into structured data tables in python. The output of this script was a row for every instance of a gene’s membership in a KEGG gene set and one column each for gene symbol, gene set (“KO reference pathway”), parent category of the gene set (“super pathway”), and the broad concept grouping of the parent category (“top-level string”). Gene symbols provided by KEGG were mapped to Ensembl identifiers with a mapping file generated using the HUGO^57^ web server on June 10th, 2020. All KEGG gene sets with the top-level string “Human” were excluded from analysis since these gene sets may be derived from studies of postmortem human tissues. Sets with less than 10 member genes were excluded from analysis, leaving 278 KEGG gene sets tested for enrichment. These 278 KEGG gene sets were tested for enrichment in 26 separate LBP gene sets: LIV DEGs, PM DEGs, the three conserved network structures across the LIV networks and PM networks (each consisting of six modules), and the six LIV PD network modules that showed at least a nominally significant overlap with the PD GWAS genes. Enrichment was tested in R using the fisher.test() function in the base stats package as the overlap between the genes in the KEGG gene set and the genes in the LBP gene set, using as background the 21,635 genes expressed in the full LBP cohort. KEGG gene sets with statistically significant associations were mapped to parent terms via the source data file. Multiple testing correction was carried out separately for each of the LBP gene sets accounting for the 278 KEGG gene sets tested using the false discovery rate estimation method of Benjamini and Hochberg^58^ implemented in R using the p.adjust() function of the base stats package.

Data used to define actively regulated genes in postmortem human brain is described in the PsychENCODE Consortium (PEC) “Integrative Flagship Paper”^21^ and was downloaded from the following:

1. High-confidence active enhancers: http://adult.psychencode.org/Datasets/Derived/DER-04b_hg38lft_high_confidence_PEC_enhancers.bed
2. Actively regulated genes: http://adult.psychencode.org/Datasets/Integrative/INT-16_HiC_EP_linkages_cross_assembly.csv

After merging these two files based on the identifiers used to define enhancers by PEC, 7,237 high-confidence active enhancers were linked to at least one of 6,324 actively regulated genes, of which 3,852 were expressed in the full LBP cohort. Fisher’s exact tests were performed between actively regulated genes and LIV-PM DEGs in R using the fisher.test() function of the base stats package using as background the 21,635 genes expressed in the full LBP cohort. Multiple testing correction was carried out to account for the 12 Fisher’s exact tests performed between enhancer bins and LIV DEGs and PM DEGs using the false discovery rate method of Benjamini and Hochberg implemented using the p.adjust() function in the base stats R package.

### DE of ALZ and SCZ performed in external postmortem human brain datasets

The datasets referred to as ALZ1 and ALZ2 throughout this report are part of the Accelerating Medicine Partnership Alzheimer’s Disease (AMPAD) initiative and have been previously described^11^. ALZ1 is the PFC sequencing data of the AMPAD Mount Sinai Brain Bank (MSBB) cohort and ALZ2 is the PFC sequencing data of the AMPAD Religious Orders Study/Memory and Aging Project (ROSMAP) cohort. For ALZ1 (N = 246 subjects), disease status was defined using the mean neocortical plaque density. For ALZ2, disease status was defined using the Consortium to Establish a Registry for Alzheimer’s Disease (CERAD) score and analyses were limited to individuals with definite ALZ (CERAD score = 1; N = 181) and no ALZ (CERAD score = 4; N = 162). ALZ1 and ALZ2 data can be downloaded from: https://www.synapse.org/#!Synapse:syn9702085

The datasets referred to as SCZ1 and SCZ2 throughout this report are part of the CommonMind Consortium (CMC) initiative and have been previously described^59^. SCZ1 refers to 96 SCZ cases and 213 controls from the National Institutes of Mental Health Human Brain Collection Core (HBCC). SCZ2 refers to 227 SCZ cases and 187 controls (after removing overlap with AMPAD) from three independent brain banks that were processed and sequenced together. SCZ1 and SCZ2 data can be downloaded from the synapse platform at http://CommonMind.org.

For each ALZ and SCZ dataset, raw PFC RNA sequencing data obtained was processed using the RAPiD pipeline as previously described^11,59^. Alignment, quantification, gene filtering, normalization, confounding technical variable identification, and sample filtering were carried out following a similar procedure described for the LBP samples. DE analyses of disease status were performed for ALZ1, ALZ2, SCZ1, and SCZ2 using the limma (v3.46.0) package in R using the following formulas:

#### ALZ1 DE

*Gene Expression ∼ Plaque Mean + Race + Sex + RIN + PMI + ExonicRate + Residuals*

#### ALZ2 DE

*Gene Expression ∼ ALZ Status + Sex + RIN + PMI + Sequencing Batch + Median 5’ to 3’ Bias + Strand Balance + Percent Intronic Bases + Residuals*

#### SCZ1 DE

*Gene Expression ∼ SCZ Status + Age at Death + Sex + PMI + RIN + IntronicRate + GenesDetected + TotalReads + IntergenicRate + Residuals*

#### SCZ2 DE

*Gene Expression ∼ SCZ Status + Age at Death + Sex + PMI + RIN + Brain Bank + ExonicRate + GenesDetected + MappedReads + IntergenicRate + Residuals*

DEGs for ALZ1, ALZ2, and SCZ2 were defined as genes with adjusted p-values < 0.05 after multiple test correction using FDR of 5%. DEGs for SCZ1 were defined as genes with unadjusted p-values < 0.05 due to the small number of genes that remained significant after p-value adjustment.

### DE of age performed in external postmortem human brain datasets

Propensity score matching was used to match the age distributions of the ALZ1, ALZ2, SCZ1, and SCZ2 datasets to the age distributions of the PM1 and PM2 datasets, respectively. Propensity score matching was implemented via the matchit() function of the MatchIt R package (version 4.1.0)^60^ with the “method” parameter set to “optimal” and the “ratio” parameter set to 1. For each of the ALZ1, ALZ2, SCZ1, and SCZ2 datasets, this resulted in two partially overlapping “age-matched” subsets – one composed of samples matched for age to the PM1 samples (“PM1 age-matched subset”) and one composed of samples matched for age to the PM2 samples (“PM2 age-matched subset”). All of the PM1 age-matched subsets consisted of 104 samples (the number of PM1 samples) and all of the PM2 age-matched subsets consisted of 126 samples (the number of PM2 samples). DE of age was performed for all PM1 age-matched subsets and all PM2 age-matched subsets using the limma (v3.46.0) and edgeR (v3.32.0)^61^ packages in R (v4.0.3) using the following formulas:

#### Age DE in PM1 age-matched subset and PM2 age-matched subset of ALZ1

*Gene Expression ∼ Plaque Mean + Age at Death + Race + Sex + RIN + PMI + ExonicRate + Residuals*

#### Age DE in PM1 age-matched subset and PM2 age-matched subset of ALZ2

*Gene Expression ∼ ALZ Status + Age at Death + Sex + RIN + PMI + Sequencing Batch + Median 5’ to 3’ Bias + Strand Balance + Percent Intronic Bases + Residuals*

#### Age DE in PM1 age-matched subset and PM2 age-matched subset of SCZ1

*Gene Expression ∼ SCZ Status + Age at Death + Sex + PMI + RIN + IntronicRate + GenesDetected + TotalReads + IntergenicRate + Residuals*

#### Age DE in PM1 age-matched subset and PM2 age-matched subset of SCZ2

*Gene Expression ∼ SCZ Status + Age at Death + Sex + PMI + RIN + Brain Bank + ExonicRate + GenesDetected + MappedReads + IntergenicRate + Residuals*

### Published DE signatures of ALZ, SCZ, BIP, and ASD

The published ALZ DE signature from AMPAD^23^ was downloaded from Synapse (syn11914808). The published SCZ, BD, and ASD DE signatures from PEC^24^ were downloaded from: https://www.science.org/doi/suppl/10.1126/science.aat8127/suppl_file/aat8127_table_s1.xlsx.

The additional 12 published ASD DE signatures from PEC^26^ were downloaded from: https://www.ncbi.nlm.nih.gov/pmc/articles/PMC9668748/bin/41586_2022_5377_MOESM5_ES M.xlsx

The published list of PD DEGs^25^ was downloaded from: https://www.ncbi.nlm.nih.gov/pmc/articles/PMC6868244/bin/41467_2019_13144_MOESM2_E SM.xlsx

### Comparing DE signatures

Spearman’s correlations between pairs of DE signatures presented throughout this report were calculated using the cor.test() function in the R base stats package. Multiple test correction for these Spearman’s correlations was not performed because the majority of unadjusted p-values returned by the cor.test() function were 0 (indicated in the main text using the “p-value < 2.2 x 10^-16^” notation). When throughout this report the overlap between two DEG sets was compared using Fisher’s exact test (e.g., LIV-PM DEGs with ALZ DEGs), the Fisher’s exact test was implemented by the fisher.test() function in the base stats R package (with the alternative parameter set to “greater”). These Fisher’s exact tests were categorized into four groups, each presented as a separate sheet in ***Supplementary Table 6***, and multiple testing correction was performed separately for each group using the false discovery rate method of Benjamini and Hochberg implemented using the p.adjust() function in the base stats R package. DE signature comparisons throughout this report were performed between DE signatures generated from non-overlapping samples when feasible (***Supplementary Table 6***). The Pearson’s correlation coefficients and associated p-values between age and PMI in the PM1 samples and PM2 samples (presented in the sub-section of the results where age DE signatures are compared) was calculated using the cor.test() function of the base stats R package.

### Correlation difference matrix analyses

Gene-gene Pearson’s correlation matrices were calculated in LIV samples and PM samples using the cor() function in the base stats R package. These calculations were performed on the residuals of the gene expression data (see “Co-expression network construction” section below). The LIV sample gene-gene correlations were subtracted from the PM sample gene-gene correlations and transformed to absolute values, resulting in the “LIV-PM correlation difference matrix.” The mean of the LIV-PM correlation difference matrix was computed. An empirical p-value was calculated to assess the significance of the LIV-PM correlation difference matrix mean. The empirical null distribution used to calculate this p-value was generated through 10,000 iterations of the following five-step procedure:

1. LIV samples were randomly split in half and PM samples were randomly split in half
2. Each of the LIV sample halves was combined with one of the PM sample halves, yielding two “random” sets of samples
3. Gene-gene Pearson’s correlation matrices were calculated for each of the random sample sets using the cor() function in the base stats R package
4. The “null correlation difference matrix” was calculated by subtracting the correlation matrix from one random set from the correlation matrix of the other random set and transforming to absolute values
5. The null correlation difference matrix mean was computed.

The empirical p-value was calculated as the fraction of 10,000 null correlation difference matrix means that were greater than the LIV-PM correlation difference matrix mean.

### Co-expression network construction

The WGCNA R package (v1.69)^55^ was used to organize genes into co-expression networks composed of functional units called modules. Residual gene expression data used in the construction of correlation matrices and co-expression networks was calculated from the full LBP cohort using Formula 10 (see above).

Six networks were constructed, each from a different subset of the full LBP cohort (i.e., LIV PD samples, LIV non-PD samples, PM1 PD samples, PM2 PD samples, PM1 non-PD samples, and PM2 non-PD samples). For each of the six subsets, samples determined to be outliers were removed. Following WGCNA convention, outliers were defined as follows:

1. Using the residual gene expression data, Euclidean distances were calculated between all pairs of samples with the dist() function of the base stats R package
2. A dendrogram was constructed through hierarchical clustering of Euclidean distances using the hclust() function of the base stats R package with the “method” parameter set to “average”
3. Outliers were labeled through manual inspection of the dendrogram.

Upon completion of outlier removal from each of the six subsets, sample sizes for each network were as follows: LIV PD network N = 194, LIV non-PD network N = 50, PM1 PD network N = 42, PM2 PD network N = 59, PM1 non-PD network N = 43, and PM2 non-PD network N = 32. Standard parameters were used to construct each network, with a soft-power threshold ranging between 6 and 8 and a minimum module size of 30 genes.

### Conserved network structure identification algorithm

To identify conserved network structures, an algorithm is necessary that considers the similarity between modules in different co-expression networks and determines whether the modules are similar enough to be considered different representations of the same biological processes. The algorithm used in this report to find conserved network structures took as input a table of “best matched” module pairs generated as follows:

1. For every network, all of the modules were compared to all of the modules in the other networks by a Fisher’s exact test implemented using the fisher.test() function in the base stats R package
2. The “best match” for a module M in a network N was defined in each of the other networks as the module with the most significant overlap with M (i.e., the module with the lowest Fisher’s exact test p-value)
3. The module pairs output by this procedure were then filtered to exclude pairs containing the grey module of any network (by WGCNA convention the grey module is not a set of co-expressed genes but rather is the set of genes that are not co-expressed with other genes).

This filtered set of best matched module pairs was then input to the conserved network structure identification algorithm. For a set of networks, *S_all_*, this algorithm is as follows:

1. One of the networks in *S_all_* is arbitrarily designated as the “reference” network (*N_ref_*). The rest of the networks are designated as the subset of “other” networks (*S_other_*) in *S_all_*. For illustrative purposes, let *S_other_* be composed of two networks, *N_other1_* and *N_other2_*
2. For each module, *M_x_*, in *N_ref_*, the following steps are performed:

a. Check whether there is a significant overlap between *M_x_* and at least one module in each of the networks in *S_other_.* If this is not the case, then *M_x_* is not a representation of a conserved network structure, and move on to consider the next module in *N_ref_*
b. If it is the case, then construct a “network-module list” that indicates the names of the networks and corresponding modules identified in the previous step (e.g., [*N_ref_*:*M_x_*, *N_other1_*:*M_y_*, *N_other2_*:*M_z_*])
c. For the modules identified in the networks of *S_other_* as the best matches for *M_x_* in *N_ref_* (i.e., *M_y_* in *N_other1_* and *M_z_* in *N_other2_*), find the best matches in *N_ref_*and in the other *S_other_* networks and construct network-module lists as was done for N_ref_
d. Compare the network-module lists to one another. If all of the lists contain the same network-module pairs, then the set of modules is considered a conserved network structure (i.e., the modules are network-specific representations of the same biological processes). In contrast, if, say, *M_q_* in *N_ref_*(rather than *M_x_*) is the best match for *M_y_* in *N_other1_*, then the network-module lists do not contain the same network-module pairs and thus this set of modules is not considered a conserved network structure
e. Check that either all or none of the modules comprising a conserved network structure are the turquoise or blue modules of their respective networks (by WGCNA convention, turquoise and blue are the two largest modules of a co-expression network). If this condition is not met, the set of modules is not considered a conserved network structure. This final step is to account for differences in module size (e.g., such that a module of 3,000 genes in one network and a module of 30 genes in another network are not considered representations of the same conserved network structure).

For some analyses presented, conserved network structures were identified in a subset of a set of networks. Let *S_all_* be a set of three networks *N_ref_*, *N_other1_*, and *N_other2_* and let *S_other_* be a subset of *S_all_* composed of *N_other1_* and *N_other2_*. Then the following steps were taken to identify “subset-specific conserved network structures” (i.e., conserved in *S_other_* but not in *S_all_*):

1. Apply the conserved network structure identification algorithm to *S_all_*. The output of this step, for example, could be [(*N_other1_*:*M_y_*, *N_other2_*:*M_z_*, *N_ref_:M_x_*)]
2. Apply the conserved network structure identification algorithm to *S_other_*. The output of this step, for example, could be [(*N_other1_*:*M_y_*, *N_other2_*:*M_z_*), (*N_other1_*:*M_v_*, *N_other2_*:*M_w_*)]
3. A subset-specific conserved network structure is then a conserved network structure in *S_other_* whose network-specific representations do not appear together in any of the conserved network structures of *S_all_*(e.g., [(*N_other1_*:*M_v_*, *N_other2_*:*M_w_*)]).

When the subset was a single network, a different procedure was followed. Let *S_all_* be a set of three networks *N_ref_*, *N_other1_*, and *N_other2_*. Let *S_other_*be a subset of *S_all_* composed only of *N_other1_*. Let *N_other1_* be composed of the modules *M_y_*, *M_v_*, and *M_t_*. Then the following steps were taken to identify subset-specific conserved network structures (i.e., conserved in *S_other_*but not in *S_all_*):

1. Apply the conserved network structure identification algorithm to *S_all_*. The output of this step, for example, could be [(*N_other1_*:*M_y_*, *N_other2_*:*M_z_*, *N_ref_:M_x_*)]
2. A subset-specific conserved network structure is then any module in *N_other1_*that is not a part of any conserved network structures identified in the previous step. The output of this step, for example, would be [(*N_other1_*:*M_v_*, *N_other1_*:*M_t_*)].

### Conserved network structure analyses

Utilizing the algorithm described in the previous section, conserved network structures were identified across the six networks built from LIV samples and PM samples. The six modules comprising each of the five conserved network structures identified were tested for enrichment of LIV DEGs and PM DEGs using Fisher’s exact test implemented using the fisher.test() function in the base stats R package. To assess the significance of the results of these tests, multiple testing correction was performed separately for the set of 30 LIV DEG enrichment tests (one test for each of the six modules comprising the five conserved network structures) and the set of 30 PM DEG enrichment tests using the false discovery rate method of Benjamini and Hochberg implemented using the p.adjust() function in the base stats R package. A conserved network structure was considered enriched for LIV DEGs or PM DEGs if over half of the six modules comprising the conserved network structure were significantly enriched after p-value adjustment. The 18 modules in the three conserved network structures enriched for LIV DEGs were tested for enrichment of KEGG gene sets as described above.

Three classes of subset-specific conserved network structures were identified for PD (“PD-specific conserved network structures”) in the networks constructed from PM samples (“PM PD networks”) and LIV samples (“LIV PD network”):

1. “LIV-PM PD structures” were defined as the subset-specific conserved network structures identified when *S_other_*was composed of the three PD networks and *S_all_* was composed of these networks plus the two PM non-PD networks.
2. “PM PD structures” were defined using the following procedure:

a. Conserved network structures were identified for the two PM PD networks
b. Subset-specific conserved network structures were identified when *S_other_*was composed of the two PM PD networks and *S_all_* was composed of these networks plus the LIV PD network
c. Subset-specific conserved network structures were identified when *S_other_*was composed of the two PM PD networks and *S_all_* was composed of these networks plus the two PM non-PD networks
d. PM PD structures were then defined as conserved network structures found in (a) whose network-specific representations did not appear together in any of the subset-specific conserved network structures found in (b) and (c)
3. “LIV PD structures” were defined using the following procedure:

a. Subset-specific conserved network structures were identified when *S_other_*was composed of the LIV PD network and *S_all_* was composed of this network plus the two PM PD networks
b. Subset-specific conserved network structures were identified when *S_other_*was composed of the LIV PD network and *S_all_* was composed of this network plus the two PM non-PD networks
c. LIV PD structures were then defined as the modules from the LIV PD network that did not appear in any of the subset-specific conserved network structures found in (a) and (b).

The three classes of PD-specific conserved network structures (LIV-PM PD structures, PM PD structures, and LIV PD structures) were tested for enrichment of PD GWAS genes using the Fisher’s exact test implemented using the fisher.test() function in the base stats R package. PD GWAS genes were defined using the default list provided on the PD GWASbrowser^62^. At the time the list was accessed (September 12th, 2021), it comprised the nearest genes to the 78 risk loci discovered in a PD GWAS of over one million individuals^31^. Gene symbols provided in the source PD GWASbrowser file were converted to Ensembl identifiers with a mapping file generated using the HUGO web server (accessed on June 10th, 2020). To assess the significance of the overlap between PD-specific conserved network structures and PD GWAS genes, multiple testing correction was performed separately for the LIV PD structures (49 tests [i.e., the number of LIV PD structures identified]), the PM1 PD network modules representing the PM PD structures (22 tests), the PM2 PD network modules representing the PM PD structures (22 tests), LIV PD network modules representing the LIV-PM PD structures (11 tests), PM1 PD network modules representing the LIV-PM PD structures (11 tests), and PM2 PD network modules representing the LIV-PM PD structures (11 tests). In each instance, the false discovery rate method of Benjamini and Hochberg was implemented using the p.adjust() function in the base stats R package. Each LIV-PM PD structure and each PM PD structure was only considered significantly enriched for PD GWAS genes if the test was significant for all of the network-specific representations of the structure. The six LIV PD structures enriched for PD GWAS genes were tested for enrichment of KEGG gene sets as described above.

## Glossary of key terms and abbreviations

● Active enhancers: enhancers located in regions of open chromatin
● Actively regulated genes: genes linked to at least one active enhancer
● ALZ: Alzheimer’s disease
● ASD: Autism spectrum disorder
● BD: Bipolar disorder
● Brain bank: Institution of origin of the PM samples (i.e., Harvard Brain Tissue Resource Center, New York Brain Bank at Columbia University, University of Miami Brain Endowment Bank)
● Co-expression network: Output of weighted gene co-expression network analysis (WGCNA); WGCNA organizes the transcriptome into modules that represent biological processes
● Co-expression network module: Group of genes in a co-expression network that show a coordinated expression pattern across samples
● Conserved network structure: In this report, a set of individual co-expression network modules (i.e., one from each separate co-expression network) considered by a rule-based algorithm to be network-specific representations of the same biological processes
● Differential expression analysis (i.e., DE): Analysis in which the expression level of every gene is tested for association with a trait of interest using a regression model; a separate regression model is fit to each gene
● Differentially expressed gene (DEG): In DE, a gene with a statistically significant association with the trait of interest (i.e., FDR-adjusted p-value for the logFC for the trait in the specific gene’s gene expression regression model is <0.05)
● DE signature: The set of logFCs for a given trait (i.e., variable) for all genes tested during differential expression analysis (i.e., in the differential expression analysis, a separate regression model that includes the trait is fit for each gene’s expression, and the DE signature is the set of all of the beta coefficients for the trait obtained from all of the gene expression regression models in the DE)
● Deep brain stimulation (DBS): Elective treatment for neurological and mental illnesses such as Parkinson’s disease (PD), obsessive-compulsive disorder, essential tremor, and dystonia
● Discovery wave cohort: 49 LIV samples and 57 PM samples sequenced in the discovery wave of RNA sequencing for the LBP
● Disease signature: DE signature when the trait of primary interest is a disease (in this report, there are disease signatures for PD, SCZ, ALZ, BD, and ASD)
● Enhancer: short DNA sequence (50-1500 base pairs in length) that regulates gene expression by serving as a binding site for transcription factors to facilitate the transcription of associated genes
● Enhancer bin: the value of a categorical variable assigned to a gene that indicates the number of high-confidence active enhancers linked to the gene (i.e., 1-5, 6-10, 11-15, 16-20, or 21-83)
● False discovery rate (FDR): In a list of statistically significant findings, the FDR is a method of p-value adjustment used to control the expected proportion of incorrectly rejected null hypotheses (“false discoveries”; features called significant that are not truly significant)
● Fisher’s exact test: A statistical test to determine if there is a non-random association between two categorical variables
● Full LBP cohort: The 518 samples (275 LIV samples and 243 PM samples) remaining after quality control procedures of the RNA sequencing data generated for this report were completed
● Gene set enrichment analysis: Method to identify whether a group of genes is over-represented or under-represented in a subset of genes as compared to the background of all genes
● Gene-gene relationship: for a pair of genes, the relative expression levels of the genes with respect to one another
● High-confidence active enhancers: active enhancers confirmed by multiple lines of evidence
● Kyoto Encyclopedia of Genes and Genomes (KEGG): Database that groups genes into sets based on known biology
● LBP: Living Brain Project
● LIV PD samples: 220 LIV samples from living participants undergoing DBS to treat PD
● LIV-PM status: Whether a PFC sample is from a living participant or a postmortem donor; the trait of primary interest in the full LBP cohort
● LIV-PM DE: DE of LIV-PM status
● LIV-PM DE signature: DE signature for LIV-PM DE
● logFC: For each gene’s regression model, the beta coefficient for a variable in the model (in this case the trait of interest), which captures both the magnitude and direction of the gene-variable association (i.e., the specific gene’s change in expression attributable to a change in the value of the variable)
● PD: Parkinson’s disease
● PEC: PsychENCODE Consortium
● PFC: Prefrontal cortex
● PM samples: Total of 246 postmortem PFC samples assembled from three brain banks
● PM PD samples: 132 PM samples from postmortem donors with a diagnosis of PD
● PM1: Total of 104 PM samples from the Harvard Brain Tissue Resource Center
● PM2: Total of 129 PM samples from New York Brain Bank at Columbia University
● PM3: Total of 13 PM samples from the University of Miami Brain Endowment Bank
● Postmortem interval (PMI): The number of hours elapsed between death and postmortem tissue processing; known confounder of postmortem brain gene expression data
● Replication wave cohort: 228 LIV samples and 187 PM samples; the samples sequenced in the replication wave of RNA sequencing for the LBP that do not overlap the samples sequenced in the discovery wave
● RIN: RNA integrity number
● SCZ: Schizophrenia
● Spearman’s correlation coefficient: Nonparametric measure used to summarize the strength and direction (negative or positive) of the relationship between two ranked variables; always between 1 and −1

## Data Availability

All data analyzed from the full LBP cohort are available via the AD Knowledge Portal (https://adknowledgeportal.org). The AD Knowledge Portal is a platform for accessing data, analyses, and tools generated by the Accelerating Medicines Partnership (AMP-AD) Target Discovery Program and other National Institute on Aging (NIA)-supported programs to enable open-science practices and accelerate translational learning. The data, analyses and tools are shared early in the research cycle without a publication embargo on secondary use. Data is available for general research use according to the following requirements for data access and data attribution (https://adknowledgeportal.org/DataAccess/Instructions). For access to content described in this manuscript see:
https://www.synapse.org/#!Synapse:syn51622714/datasets/

## Acknowledgments

The Living Brain Project study participants are commended for their important role in science. The following centers, programs, departments and institutes within the Icahn School of Medicine at Mount Sinai supported the work: Department of Neurosurgery; Department of Genetics and Genomic Sciences; Department of Psychiatry; Department of Neuroscience; Department of Medicine; Friedman Brain Institute; Department of Anesthesia; Department of Pathology. Postmortem samples from Harvard Brain Tissue Resource Center and University of Miami Brain Endowment Brain were acquired under the National Institutes of Health NeuroBioBank request number 543. Postmortem samples from the New York Brain Bank of Columbia University were acquired under request number 1962. The datasets referred to as SCZ1 and SCZ2 throughout this report are part of the CommonMind Consortium initiative and were utilized under the National Institutes of Mental Health Repository and Genomics Resource request identifier 5bffe32e97024. The datasets referred to as ALZ1 and ALZ2 throughout this report are part of the Accelerating Medicine Partnership Alzheimer’s Disease initiative. ALZ1 data were generated from postmortem brain tissue collected through the Mount Sinai VA Medical Center Brain Bank and were provided by Dr. Eric Schadt from the Icahn School of Medicine at Mount Sinai. ALZ1 data were generated from postmortem brain tissue collected through the Mount Sinai VA Medical Center Brain Bank and were provided by Dr. Eric Schadt from the Icahn School of Medicine at Mount Sinai. ALZ2 data were generated from postmortem brain tissue collected through the Rush Alzheimer’s Disease Center, Rush University Medical Center, Chicago, and data collection was supported through funding by NIA grant R01AG15819 (ROSMAP; genomics and RNAseq). ALZ1 and ALZ2 data were accessed utilizing the Alzheimer’s Disease Knowledge Portal, which would not be possible without the participation of research volunteers and the contribution of data by collaborating researchers. The following individuals are acknowledged for their advice and feedback over the course of the study: Elenita Sambat, Douglas M. Ruderfer, Wayne Goodman, Milind Mahajan, Eric J. Nestler, and Dennis S. Charney.

## Data and materials availability

All data analyzed from the full LBP cohort are available via the AD Knowledge Portal (https://adknowledgeportal.org). The AD Knowledge Portal is a platform for accessing data, analyses, and tools generated by the Accelerating Medicines Partnership (AMP-AD) Target Discovery Program and other National Institute on Aging (NIA)-supported programs to enable open-science practices and accelerate translational learning. The data, analyses and tools are shared early in the research cycle without a publication embargo on secondary use. Data is available for general research use according to the following requirements for data access and data attribution (https://adknowledgeportal.org/DataAccess/Instructions). For access to content described in this manuscript see: https://www.synapse.org/#!Synapse:syn51622714/datasets/

